# Long COVID Brain Fog Treatment: Findings from a Pilot Randomized Controlled Trial of Constraint-Induced Cognitive Therapy

**DOI:** 10.1101/2024.07.04.24309908

**Authors:** Gitendra Uswatte, Edward Taub, Karlene Ball, Brandon S. Mitchell, Jason A. Blake, Staci McKay, Fedora Biney, Olesya Iosipchuk, Piper Hempfling, Elise Harris, Anne Dickerson, Kristine Lokken, Amy J. Knight, Victor W. Mark, Shruti Agnihotri, Gary Cutter

## Abstract

**Purpose:** Long COVID brain fog is often disabling. Yet, no empirically-supported treatments exist. This study’s objectives were to evaluate feasibility and efficacy, provisionally, of a new rehabilitation approach, Constraint-Induced Cognitive Therapy (CICT), for post-COVID-19 cognitive sequelae.

**Design:** Sixteen community-residents ≥ 3-months post-COVID-19 infection with mild cognitive impairment and dysfunction in instrumental activities of daily living (IADL) were enrolled. Participants were randomized to Immediate-CICT or treatment-as-usual (TAU) with crossover to CICT. CICT combined behavior change techniques modified from Constraint-Induced Movement Therapy with Speed of Processing Training, a computerized cognitive-training program. CICT was deemed feasible if (a) ≥80% of participants completed treatment, (b) the same found treatment highly satisfying and at most moderately difficult, and (c) <2 study-related, serious adverse-events occurred. The primary outcome was IADL performance in daily life (Canadian Occupational Performance Measure). Employment status and brain fog (Mental Clutter Scale) were also assessed.

**Results:** Fourteen completed Immediate-CICT (*n*=7) or TAU (*n*=7); two withdrew from TAU before their second testing session. Completers were [*M* (S*D*)]: 10 (7) months post-COVID; 51 (13) years old; 10 females, 4 males; 1 African American, 13 European American. All the feasibility benchmarks were met. Immediate-CICT, relative to TAU, produced very large improvements in IADL performance (*M*=3.7 points, p<.001, *d*=2.6) and brain fog (*M*=−4 points, p<.001, *d*=−2.9). Four of five non-retired Immediate-CICT participants returned-to-work post-treatment; no TAU participants did, *p*=.048.

**Conclusions:** CICT has promise for reducing brain fog, improving IADL, and promoting returning-to-work in adults with Long COVID. Findings warrant a large-scale RCT with an active-comparison group.

**IMPACT:**

- Brain fog in adults with Long COVID is often associated with dysfunction in everyday activities and unemployment. Yet, there are no empirically supported treatments targeting cognition in this population. Findings from this small-scale, pilot randomized controlled trial (RCT) suggest that a novel intervention, i.e., Constraint-Induced Cognitive Therapy, is a feasible cognitive rehabilitation method in adults with Long COVID cognitive sequelae with promise of (a) improving performance of cognition-based tasks in daily life and (b) promoting return-to-work. Further studies with larger sample sizes are warranted.
- Speed of Processing Training (SOPT) has been shown to increase processing speed in older adults without neurological disorders but has not been applied to adults with brain fog due to Long COVID, in whom slowing of cognitive processing speed is common. The results of this pilot RCT suggest that SOPT, in conjunction with behavior change techniques, may increase cognitive processing speed in this brain-injured population.

## INTRODUCTION

The worldwide spread of coronavirus disease 2019 (COVID-19) represents the largest pandemic since influenza B in 1935.^1^ Current estimates from the U.S. Centers for Disease Control and Prevention suggest that about 11% of those who contract COVID-19 develop chronic symptoms, i.e., “long COVID” or Post-Acute Sequelae of SARS-COV-2 (PASC).^2^ The symptoms include “brain fog” and cognitive impairment, fatigue, anxiety, depression, and shortness of breath and other physical problems.^3,4^ Increasing evidence suggests that CNS inflammation, along with microvascular and cellular damage, contribute to the neuropsychological symptoms.^5^

Brain fog, which is the experience of confusion, forgetfulness, and sluggish thinking,^6,7^ and cognitive dysfunction are among the most common PASC symptoms.^3^ In a 56-country 2020 Internet survey (*N*=3,762), 85% of adults with PASC endorsed these two symptoms.^8^ In a U.S., nonprobability, population-based 2023 Internet survey (N=14,767), 57% of adults with PASC reported difficulty with at least one of the following: slowed thinking, decision-making, multi-tasking, memory, starting tasks, attention, and concentration.^9^ These perceptions are accompanied typically by mild impairments on neuropsychological tests of processing speed, executive function, memory encoding and recall, and phonemic and category fluency.^10^ Brain fog, however, remains disabling in many. Adults with PASC are less likely to have full-time jobs and more likely to be unemployed than before COVID-19 infection^11^ and report brain fog as the main cause of difficulties with work duties such as remembering routine tasks, learning new tasks, and communicating with others.^12^ Systematic reviews show that adults with PASC also have difficulties with performing everyday tasks with important cognitive components, i.e., instrumental activities of daily living (IADL).^13^ Yet, there are no interventions for brain fog and cognitive dysfunction in this population with any evidence for efficacy from a randomized controlled trial (RCT).^14^

Constraint-Induced Cognitive Therapy (CICT) is a new rehabilitation method that our laboratory has applied to stroke survivors with mild-to-moderate cognitive impairment with promising results.^15^ CICT combines two interventions: Speed of Processing Training (SOPT)^16,17^ and a modified version of the Transfer Package of Constraint-Induced Movement Therapy (CIMT)^18–20^ focused on cognition. For both interventions, efficacy is supported by multiple, single-site, randomized controlled trials (RCT) and several large multisite RCTs (SOPT,^16,17^ CIMT^18–20^). SOPT is computerized cognitive training that requires users to identify and locate targets on a monitor; cognitive load is increased as the user progresses by, for example, adding distractors.^17^ Results from the largest-to-date RCT of cognitive interventions in community-dwelling older adults indicate that SOPT produces long-lasting benefits on in-lab tests of (a) cognitive processing speed^16,21^ and (b) IADL performance.^22,23^ Benefits are also present in improved driving in the real world.^24^ However, SOPT’s impact on other cognition-based IADL outside of the lab is mixed.^15^ The Transfer Package contains behavior change techniques designed to transfer gains from the treatment setting to daily life.^25,26^ **Figure 1** sketches the mechanisms by which we think CICT operates.

**Figure 1.**
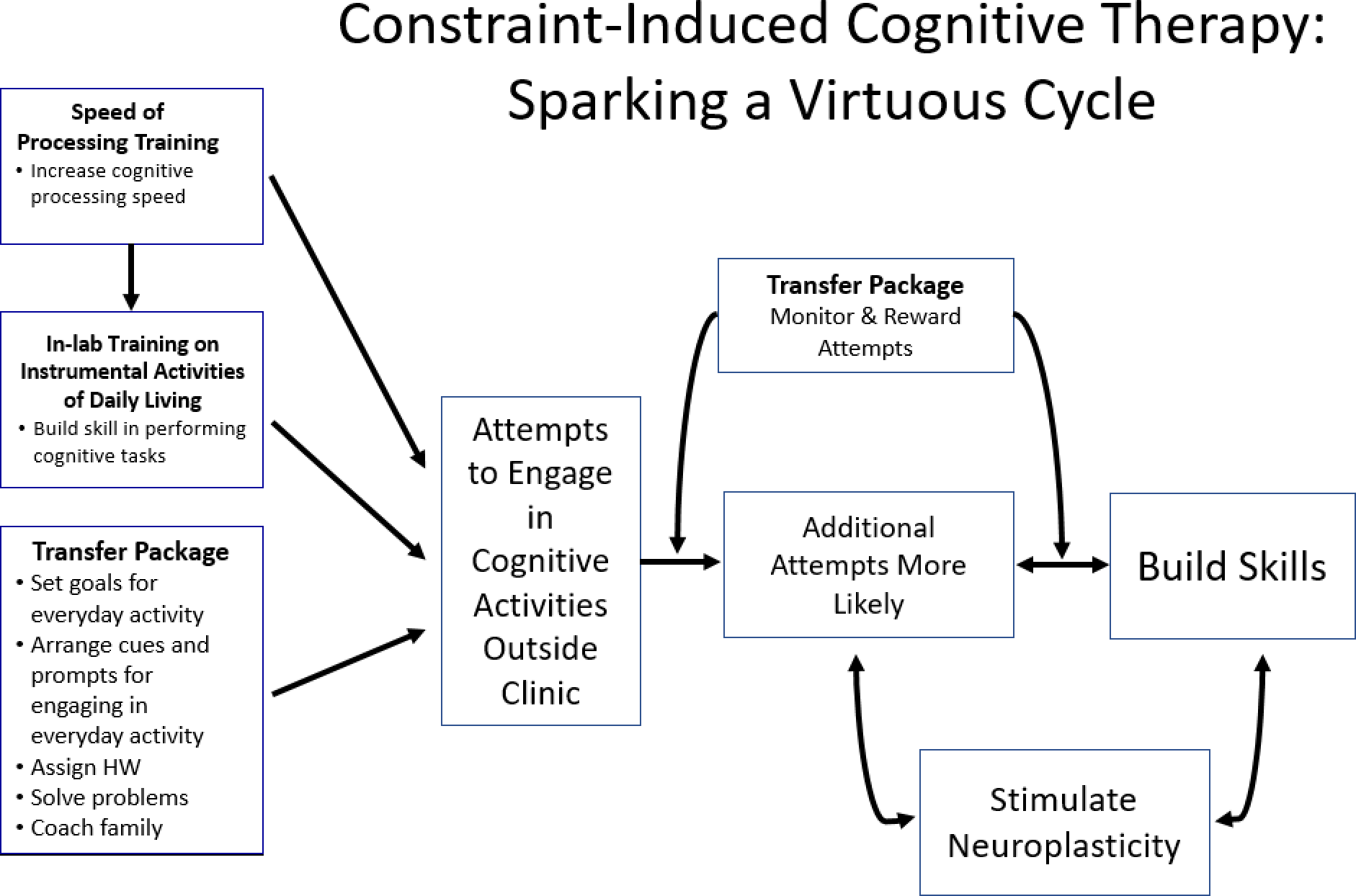
A Conceptual Model of How Constraint-Induced Cognitive Therapy (CICT) Operates. The three components of CICT are listed in the boxes on the left-hand side of the figure. We hypothesize that all three components promote attempts to perform cognition-based tasks outside the treatment setting. Moving from left to right, we hypothesize elements of the Transfer Package (i.e., the Cognitive Task Activity Log, Inventory of New Cognitive Activities, and Home Skill Assignment; see **Supplement 1**, **Table s2**) permit therapists to monitor and reward attempts when they occur.^50^ Rewarding behavior increases its frequency.^31,32^ Repetition builds skill and stimulates brain plasticity.^51^ Skill building and neuroplasticity support each other,^52^ which we hypothesize, in turn, makes attempts at cognition-based tasks less effortful and thereby more frequent—kicking off a virtuous cycle.^50^

The large improvements observed in our stroke pilot, along with the overlap in stroke and PASC neuropathology and cognitive symptoms,^27^ including reduced cognitive processing speed,^28,29^ prompted us to test CICT in post-COVID adults with persistent brain fog accompanied by mild cognitive impairment and IADL dysfunction. The pilot RCT described herein aims to evaluate the feasibility and efficacy, on a preliminary basis, of CICT for rehabilitating everyday cognitive function in this population.

## METHODS

### Study Design

Participants in this pilot RCT with an unblinded, open-label, parallel arm, partial-crossover design, were randomized in blocks of two by the project coordinator to receive CICT immediately or any treatment-as-usual (TAU) from healthcare providers. TAU participants were crossed over to CICT three months afterwards (**Supplement 1, Figure s1**). Random assignment was performed using a computer-generated random-numbers table, which the project data manager set up. In addition to assessing feasibility, outcomes were evaluated, on a preliminary basis, in three domains: everyday cognitive task performance, psychological distress, and in-lab cognitive ability. The primary endpoint was pre-to post-treatment change on the Canadian Occupational Performance Measure Performance Scale (see **Measures**). For Immediate-CICT participants, testing occurred before and after treatment. For TAU participants, testing occurred on parallel occasions (Baseline 1 & 2) during the TAU period and before and after crossover to CICT.

### Participants

Adults ≥3 months from their initial COVID-19 infection of any severity with brain fog symptoms were recruited from the University of Alabama at Birmingham post-COVID clinic. In addition, a few candidates made contact after press reports about the lab’s work. Inclusion criteria included mild to moderate cognitive impairment per a Montreal Cognitive Assessment^30^ score between 10-26 and some impairment in IADL per a Cognitive Task Activity Log score ≤3.5 (see **Supplement 1**). Participants had to be community residents, have reliable transportation, be medically stable, and have adequate sight and hearing to complete testing. Individuals with pre-existing cognitive impairment, such as those with dementia, traumatic brain injury and stroke, were excluded, as were those with severe depression or frailty. Enrollment, which occurred from January 2021 to August 2022, was performed by the screeners.

### Intervention: Constraint-Induced Cognitive Therapy (CICT)

Thirty-six hours of training were scheduled over 2 to 7 weeks depending on participants’ needs (**Table 1**). Each training session included SOPT (approximately 20%), in-lab training on IADL (35%), the Transfer Package (30%), and rest periods (15%). Each treatment component is described briefly below and at greater length in **Supplement 1**.

**Table 1.** Participant Demographic, COVID-19, and CICT Treatment Characteristics.

| Characteristic | Immediate-CICT<br>(n=7) | TAU<br>(n=7) | All<br>(N=14) |
| --- | --- | --- | --- |
| Age, years | 45 (15.2) | 56.4 (6.9) | 50.7 (12.8) |
| Sex |  |  |  |
| Male | 1 | 3 | 4 |
| Female | 6 | 4 | 10 |
| Ethnicity |  |  |  |
| European American | 6 | 7 | 13 |
| African American | 1 | 0 | 1 |
| Education | 15.7 (2.3) | 15.2 (2.3)<sup>a</sup> | 15.5 (2.2) |
| Months since COVID-19 onset | 7.4 (2) | 12.9 (7) | 10.1 (6.5) |
| Hospitalized due to COVID-19 | 0 | 2 | 2 |
| CICT training duration |  |  |  |
| Hours, *M* (*SD*), min-max | 34.8 (3.1), 30.8-39.8 | 32.2 (4.6), 26.5-35.8<sup>b</sup> | 33.8 (3.7), 26.5-39.8 |
| Sessions, *M* (*SD*), min-max | 12 (1.8), 9-15 | 11 (1), 10-12<sup>b</sup> | 11.5 (1.6), 9-15 |
| Days from first to last session, *M* (*SD*), min-max | 27 (13.5), 10-52 | 31.3 (12.5), 15-41<sup>b</sup> | 28.6 (12.7), 10-52 |

Abbreviations: CICT, Constraint-Induced Cognitive Therapy; TAU, treatment-as-usual.
Values are *M* (*SD*) or counts. There were no significant between-group differences for any characteristics.
<sup>a</sup> Years of education were missing for two TAU participants.
<sup>b</sup> Three TAU participants dropped out before crossover to CICT. Hence, crossover-CICT data were only available for four.

The *SOPT* software requires that participants rapidly detect, identify, discriminate, and locate targets on a monitor in successive exercises, each more difficult than the previous one. On the first exercise, for example, participants are asked to identify a target at the center of the monitor. On the second exercise, participants are asked to identify a central target and locate another target in the periphery. In addition, the target display time is decreased on each exercise as participants improve.^16,17, 23^

*In-lab IADL Training*, along with the Transfer Package, is designed to bridge SOPT to performance of IADL outside the lab. Participants receive training on IADL following shaping principles, which involve approaching a behavioral objective in small steps with training chunked into brief, readily quantifiable, trials and provision of frequent positive reinforcement.^31–33^ Task practice, which employs continuous, less easily quantifiable, tasks, was also used.^18,20^ Examples of tasks are generating a shopping list, making an appointment calendar, and drafting a work email.

The *Transfer Package* here was a close analog for cognitive tasks of the Motor Transfer Package used in the studies described in the **Introduction**. **Supplement 1, Table s2** lists its elements.^25,26^

### Feasibility

Adherence to, engagement with, and acceptance of CICT by participants were measured. Adherence was quantified by the number of treatment hours and homework tasks completed. Engagement was indexed by (1) the number of everyday tasks resumed after starting treatment, which was measured with an in-house behavior log, the Inventory of New Cognitive Activities (INCA),^15^ and (2) changes in how independently and how well everyday tasks were performed, which were measured with an in-house, structured, patient-centered interview, the Cognitive Task Activity Log (CTAL).^15^ Acceptance was assessed using an in-house survey, i.e., the Participant Opinion Survey (POS), featuring 7-point scales that quantify satisfaction with, perception of benefit from, and difficulty of the intervention. The INCA, CTAL, and POS are described in **Supplement 1**. Safety was monitored by logging adverse events in consultation with the project Medical Director (VWM).

### Outcomes

*Canadian Occupational Performance Measure* (COPM). This validated, widely-used, patient-centered, transdiagnostic, structured interview has been used to measure changes in self-rated occupational performance (e.g., self-care, productivity, and leisure) over time.^34,35^ Here, five, self-selected activities with important cognitive components were rated on performance quality (Performance scale; 1=not able, 10=able to do it extremely well) and satisfaction (Satisfaction scale; 1=not at all, 10=extremely). A minimal clinically important difference (MCID) is 2 on each of these 10-point scales.^36^

*Employment status*. Employment status was assessed at three points, (1) before COVID onset, (2) after COVID onset, (3) after CICT. Participants were assigned into one of the following categories: employed, unemployed, and retired. Participants also reported where they worked and whether they were able to fulfill their duties.

*Mental Clutter Scale* (MCS). This 8-item self-report scale assesses the severity of brain fog symptoms. Respondents rate how frequently they experience 8 symptoms (e.g., fuzzy-headedness, cluttered thinking) using a 10-point scale (1=not at all, 10=all the time).^37^

*Fatigue Assessment Scale (FAS).* This self-report scale quantifies how frequently respondents experience ten fatigue symptoms with a 5-point scale (1=never, 5=always).^38^ The FAS MCID is 4.^39^

The *Patient Health Questionnaire-9*^40^ (PHQ-9, max=27, MCID=5)^41^ assesses depressive symptom frequency; the *General Anxiety Disorder-7*,^42^ (GAD-7, max=21, MCID=4)^43^ assesses anxiety symptom frequency. Both are standard transdiagnostic measures.^40–43^

*Symbol Digit Modalities Test - Oral Version*: The *SDMT* is a standard, transdiagnostic measure of information processing speed.^44,45^ Participants are shown an array of abstract symbols along with a key pairing each unique symbol with a number from 1-9; participants are asked to say the number for each symbol in the array. The score is the number of symbols coded correctly in 90 seconds (max=110; MCID=4).^46^

*Montreal Cognitive Assessment (MoCA)*. This standard, transdiagnostic, cognitive screen assesses a broad array of cognitive functions^30^ (max=30, MCID=2).^47^

### Data Analysis

Power calculations were not performed to determine a sample size sufficient to reliably detect statistically significant changes for this pilot. Its primary purpose was to evaluate CICT’s feasibility, which was done by calculating whether ≥80% of participants met the following benchmarks: (1) completed ≥80% of treatment hours prescribed, (2) completed ≥70% of homework assigned, (3) found CICT highly satisfying (≥6 on relevant POS item), (4) found CICT highly beneficial (≥6 on relevant POS item), and (5) found CICT to be at most moderately difficult (≤5 on relevant POS item). A sixth benchmark was ≤2 study-related, serious adverse-events.

CICT’s efficacy was evaluated on a preliminary basis by using analysis of covariance (ANCOVA). Separate models, which adjusted for baseline scores, were used to compare scores after CICT and TAU on each of the outcomes except employment. Effect sizes were described using Cohen’s *d*; values ≥0.8 are large.^48^ Non-parametric ANCOVAs were used to analyze the COPM Satisfaction, MoCA, SDMT, FAS, and PHQ-9 data because they deviated from normality per review of Q-Q plots, outliers, and Shapiro-Wilk test statistics. For these non-parametric models, ranks were substituted for raw scores. Raw-score statistics are reported for the COPM Satisfaction scale, MoCA, and FAS because the non-parametric and parametric models produced similar results. Dissimilar results were observed for the PHQ-9 and SDMT; hence, rank-based and raw-score statistics are reported in the **Results** and **Supplement 1**, respectively. Fisher’s Exact Test was used to compare employment status after CICT vs TAU. Analyses of changes in the TAU group after crossover to CICT are described in **Supplement 1**. All data are reported on a completers basis because the completers analyses were more conservative than the intention-to-treat analyses; all the drop-out occurred in the TAU group (see below). We did not correct for multiple comparisons because our focus was on the feasibility endpoints. However, all nine outcomes showed an advantage for the same group; the probability of that occurring by chance was only 0.2%. All analyses were performed using IBM SPSS.

### Transparency and Openness

The study was approved by the Institutional Review Board at the University of Alabama at Birmingham; all participants gave written informed consent. The study’s design was pre-registered; see clinicaltrials.gov/study/NCT04644172. The study write-up follows the Consolidated Standards of Reporting Trials (CONSORT) RCT checklist. All data exclusions and manipulations are reported. The study materials, de-identified data, and analytic code are available by emailing the corresponding author.

## RESULTS

### Participant Characteristics

**Supplement 1, Figure s1** depicts participants’ flow through the study. Seven were assigned to Immediate-CICT; nine to TAU. Two assigned to TAU withdrew before Baseline 2 testing: one lost interest in the study, the other had transportation problems. They have been excluded from all the analyses. An additional three TAU participants dropped out prior to crossover to CICT because of medical problems (*n*=2) or loss of interest (*n*=1); they have been excluded from the feasibility calculations below and crossover descriptive statistics in **Supplement 1**. Due to scheduling issues, one of these participants did not complete the COPM, MCS, and SDMT at Baseline 1; in addition, his Baseline 1 MoCA score was an outlier (**Supplement 1, Table s1**). Hence, he was excluded from the efficacy analyses of these measures. Although TAU participants were permitted to receive treatment from healthcare providers in the community before crossover to CICT, none received any therapy for PASC cognitive sequelae. **Table 1** lists participants’ demographics. There were no significant differences at baseline between Immediate-CICT and TAU participants’ age, PASC chronicity, gender, education, and unemployment; neither were there on the outcomes.

### Feasibility

All Immediate- and Crossover-CICT participants but one adhered to the requirement to complete ≥29 hours of CICT (mean [SD]= 33.8 [3.7]; **Table 1**); one Crossover-CICT participant did only 26.5 hours because of scheduling conflicts. All but one met the threshold for adhering to the homework (80% [17.5%]; **Table 1**); one Immediate-CICT participant completed only 37% of her assignments. Six Immediate-CICT participants met the thresholds on the POS for satisfaction with CICT (7 [0] points) and perceived benefit from CICT (6.8 [0.4] points). POS data were missing from one but her family caregiver perceived benefit that was high (6 out of 7). Five other participants’ family caregivers also perceived high benefit (6.5 [0.5]). One participant did not have a family caregiver. Immediate-CICT participants reported that CICT was only moderately difficult (3.6 [2.04] points). Crossover-CICT participants were excluded from the description of CICT’s acceptability because POS data were available for only two. The INCA and CTAL data, which index engagement, are in **Supplement 1**. There were no study-related adverse events.

### Efficacy: Everyday Activities and Employment

A very large advantage in favor of Immediate-CICT over TAU was observed after treatment on the primary outcome: COPM Performance scale mean difference^49^ (*MD*)=3.7 points; *95% CI*, 2.5-4.9; *F*(1,10)=51, *p*<.001; *d*=2.6 (**Figure 2**). An interaction effect, which is reported in **Supplement 1**, was present on the COPM Satisfaction scale. Post-crossover COPM changes in TAU participants, along with post-crossover changes on all the other outcomes, are described in **Supplement 1**.

**Figure 2.**
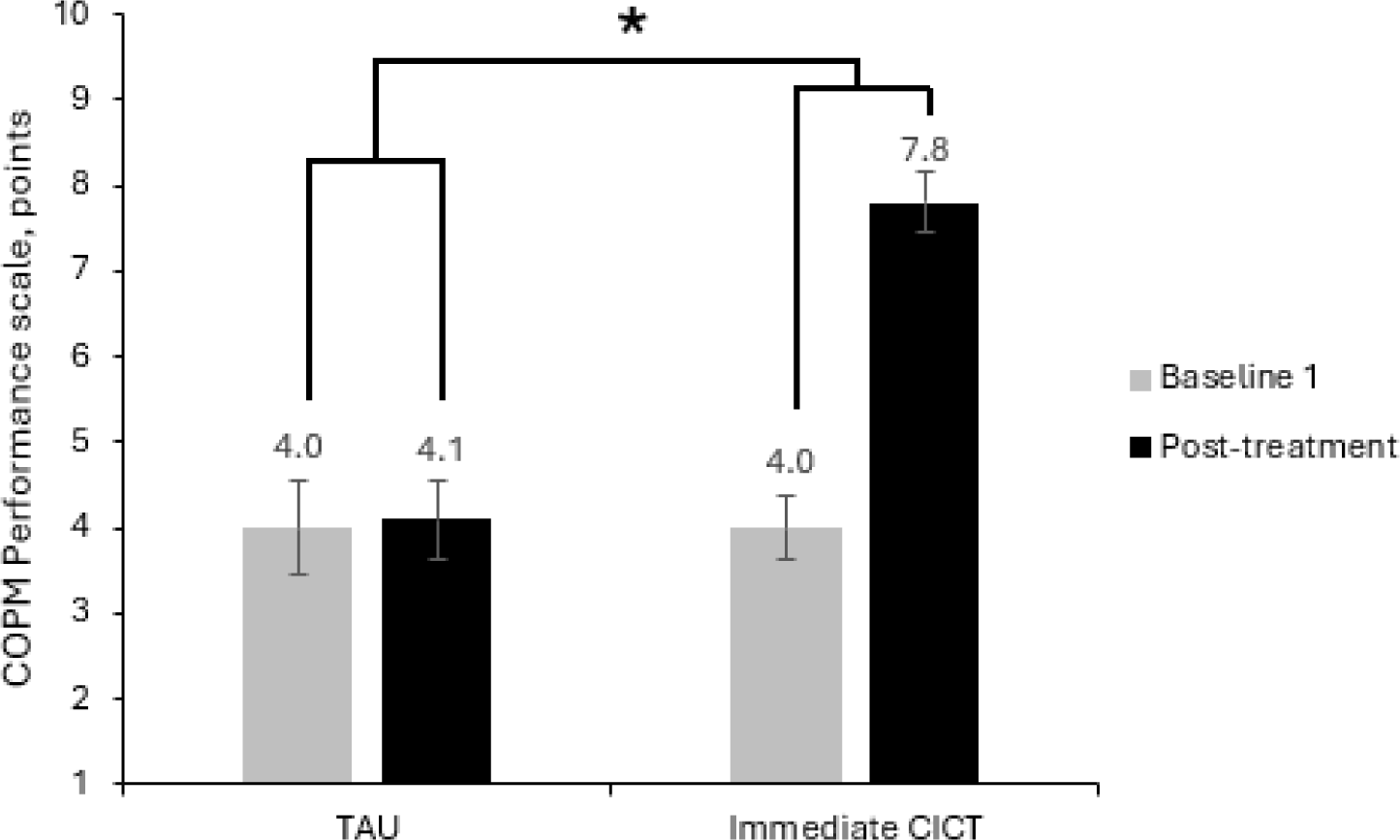
Performance of Everyday, Cognition-based Activities Before and After Immediate-Constraint-Induced Cognitive Therapy (CICT) and treatment-as-usual (TAU). The Canadian Occupational Performance Measure (COPM) Performance scale measures how well participants perform five self-selected cognition-based activities (1 = not able; 10 = able to do it extremely well). Horizontal bars represent standard errors. All Immediate-CICT participants had clinically meaningful improvements; no TAU participants did. \**p* < *.001*

A distinct advantage in favor of Immediate-CICT over TAU was also observed in return-to-work, *p*=0.048 (**Table 2**). In both groups, two had retired prior to COVID-19 onset. Out of the remaining five Immediate-CICT participants, four had to give up their job after COVID-19 onset, and one switched to remote-work, fulfilling only a limited duty set. After Immediate-CICT, four of five were able to resume work with a full duty set. None were able to work before or after TAU.

**Table 2.**
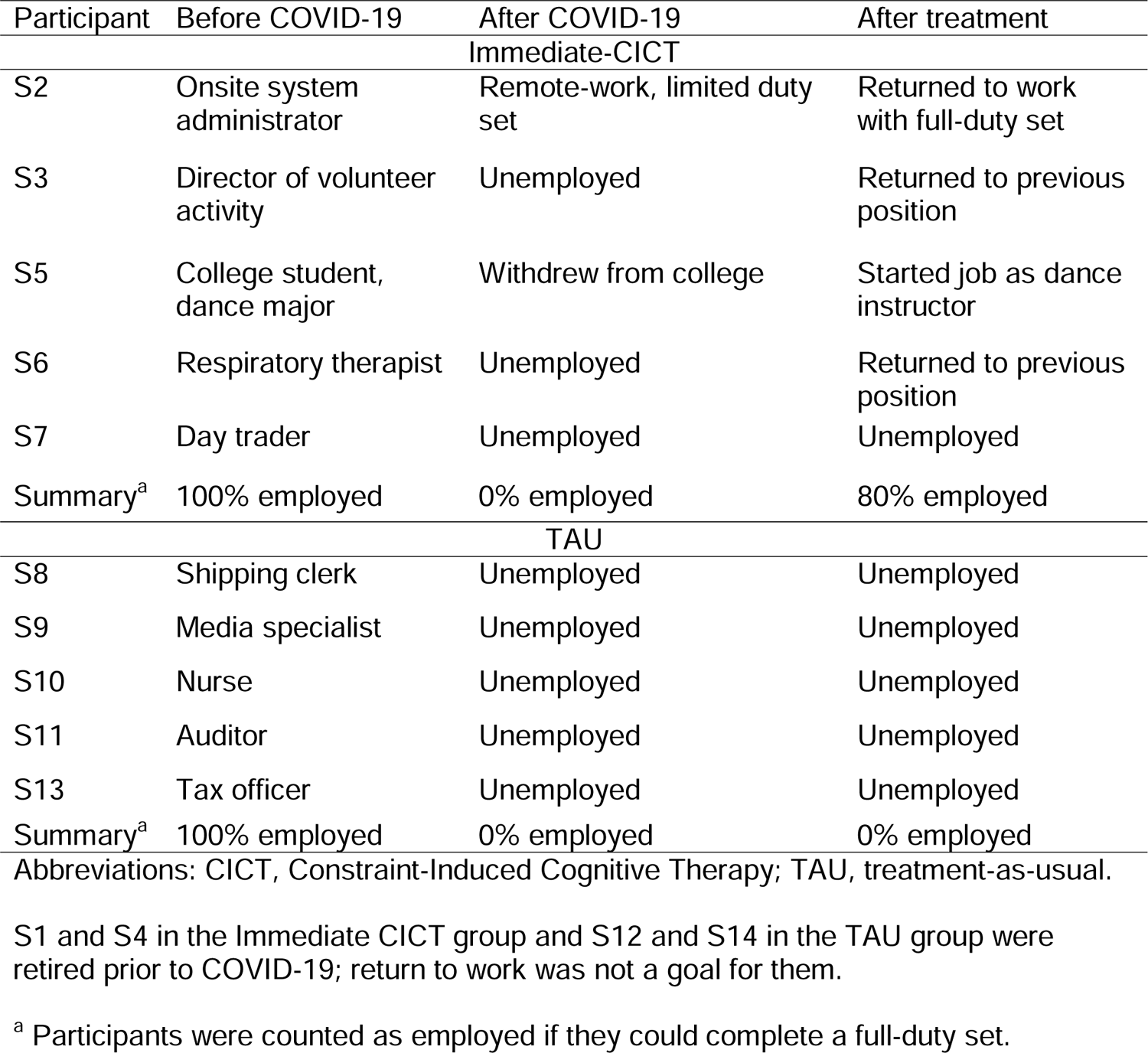
Participants’ Employment Status Before and After COVID-19 Onset and After Treatment.

| Participant | Before COVID-19 | After COVID-19 | After treatment |
| --- | --- | --- | --- |
| Immediate-CICT |  |  |  |
| S2 | Onsite system administrator | Remote-work, limited duty set | Returned to work with full-duty set |
| S3 | Director of volunteer activity | Unemployed | Returned to previous position |
| S5 | College student, dance major | Withdrew from college | Started job as dance instructor |
| S6 | Respiratory therapist | Unemployed | Returned to previous position |
| S7 | Day trader | Unemployed | Unemployed |
| Summary<sup>a</sup> | 100% employed | 0% employed | 80% employed |
| TAU |  |  |  |
| S8 | Shipping clerk | Unemployed | Unemployed |
| S9 | Media specialist | Unemployed | Unemployed |
| S10 | Nurse | Unemployed | Unemployed |
| S11 | Auditor | Unemployed | Unemployed |
| S13 | Tax officer | Unemployed | Unemployed |
| Summary<sup>a</sup> | 100% employed | 0% employed | 0% employed |

Abbreviations: CICT, Constraint-Induced Cognitive Therapy; TAU, treatment-as-usual.
S1 and S4 in the Immediate CICT group and S12 and S14 in the TAU group were retired prior to COVID-19; return to work was not a goal for them.
<sup>a</sup> Participants were counted as employed if they could complete a full-duty set.

### Efficacy: Psychological Distress

Immediate-CICT, compared to TAU, resulted in very large reductions in brain fog symptoms (MCS *MD*=−4 points; *95% CI*, −5.3 to −2.6.; *F*[1,10]=43, *p*<0.001; *d*=−3.1; **Figure 3**) and fatigue (FAS *MD*=−10.9 points; *95% CI*, −17.4 to −4.4; *F*[1,10]=7.6, *p*=0.02; *d*=−1.8). For the latter, the advantage of CICT over TAU was larger for participants with high baseline scores than for participants with low scores (**Supplement 1**). A large benefit from Immediate-CICT, relative to TAU, was observed for depressive symptoms, *F*(1,11)=7.5, *p*=.019 (**Supplement 1, Figure s12**). The median post-treatment PHQ-9 score in the Immediate-CICT group after treatment was 6, inter-quartile range (IQR)=5-7; the corresponding value in the TAU group was 10.0, IQR=7-11. Although an advantage was observed for Immediate-CICT over TAU after treatment in anxiety symptoms, the difference was not statistically significant: GAD-7 *MD*=−3.3 points; *95% CI*, −7.3 to 0.6; *F*(1,11)=3.4, *p*=0.09; *d*=−0.8.

**Figure 3.**
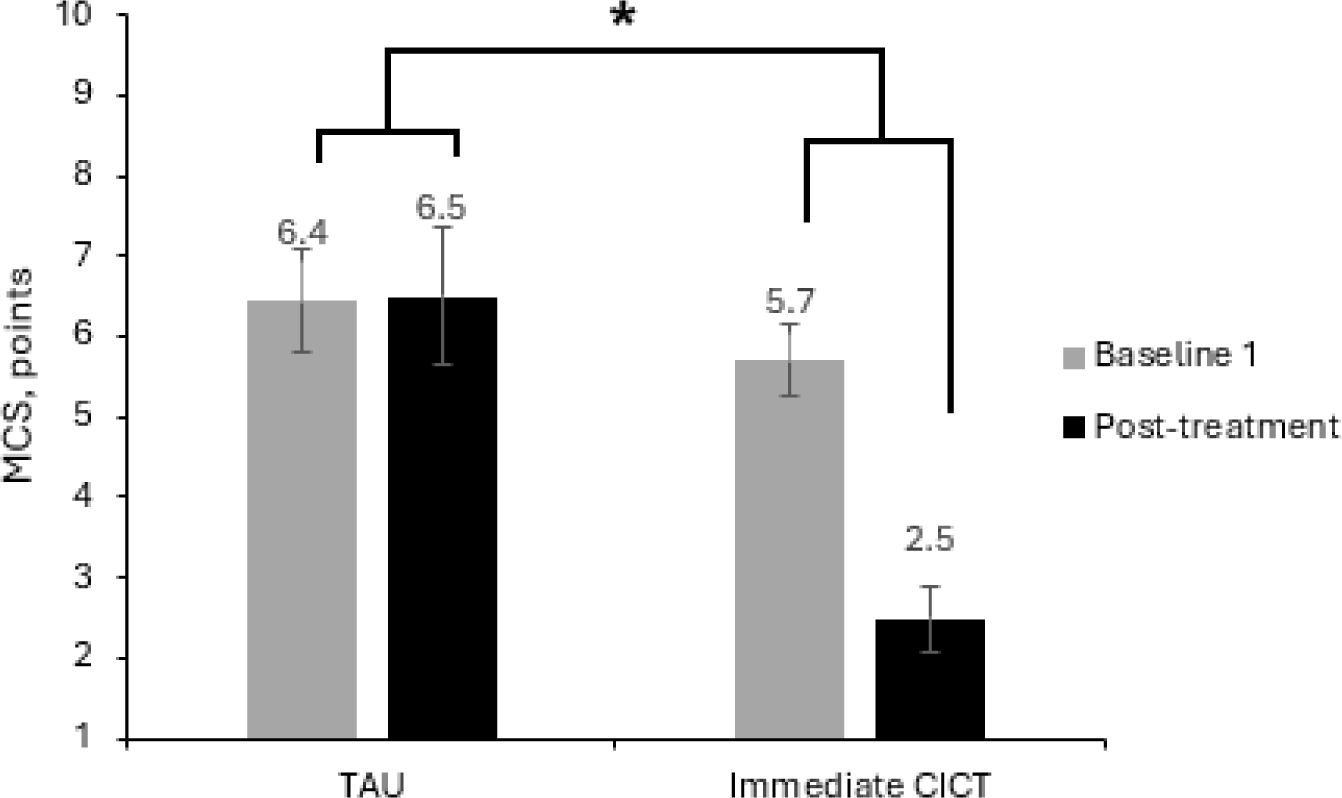
Brain Fog Symptom Frequency Before and After Immediate-Constraint Induced Cognitive Therapy (CICT) and treatment-as-usual (TAU). The Mental Clutter Scale (MCS) measures how frequently participants experience eight brain fog symptoms (1 = not at all, 10 = all the time). Horizontal bars represent standard errors. *\*p* < *.001*

### Efficacy: In-lab Cognitive Testing

A moderate benefit from Immediate-CICT, relative to TAU, was observed for cognitive processing speed, *F*(1,10)=5.4, *p*=0.042 (**Supplement 1, Figure s13**). The median post-treatment SMDT score in the Immediate-CICT group was 51, IQR=29-56; the corresponding value in the TAU group was 39, IQR=37-49. No advantage for Immediate-CICT over TAU was seen for general cognitive ability: MoCA *MD*=0.7 points; *95% CI*, −3.4-4.8; *F*(1,10)=0.1, *p*=0.73; *d*=0.2 (**Supplement 1, Table s1**).

## DISCUSSION

The results suggested that CICT is a feasible method for reducing disability in adults with brain fog and cognitive dysfunction due to PASC; all the benchmarks for adherence, acceptability, and safety were met. Moreover, CICT, compared to TAU, resulted in very large improvements in performance of cognition-based activities in daily life. All Immediate-CICT participants reported clinically meaningful improvements on the COPM Performance scale; no TAU participants did so. Skeptics might argue the substantial advantage in favor of CICT on the primary outcome was due to the operation of demand characteristics, e.g., the desire of CICT participants to please experimenters. However, the changes in employment observed suggest otherwise: 80% of Immediate-CICT participants who had not retired prior to COVID-19 onset resumed a full set of work duties after treatment; none did so after TAU. Even though employment was assessed by self-report, return-to-work might be considered a “hard” endpoint because of its binary nature and description of a state-of-the-world (as opposed to an internal state). An advantage for CICT over TAU was also observed on the SDMT, which is an objective cognitive-processing-speed test.

Immediate-CICT participants, compared to TAU participants, reported very large reductions in brain fog and fatigue and large reductions in depressive symptoms. One interpretation of the results is that CICT benefited IADL, employment, and brain fog by improving participants’ mood. Another is that CICT produced improvements in both sets of variables by targeting a common mechanism or targeting multiple mechanisms. Regardless, CICT produced improvements in both everyday function and psychological distress.

### Constraints on Generality and Other Study Limitations

The small (*n*=14) and ethnically homogenous (93% European American) sample raises questions about the generality of the findings. Future trials might consider tailoring their recruitment strategies to ethnic groups who are under-represented here. The preponderance of females (*n*=10) in the sample is typical of the target population;^3,9^ females appear to be at higher risk of developing Long COVID because of unique features of their immune system.^3^ Other important limitations were the absence of long-term follow-up, blinding, and control for placebo effects. A shortcoming of the cognitive outcome assessment was use of the MoCA, which was designed for use as a screening tool.^30^ Two-of-nine TAU participants withdrew before Baseline 2 testing, which may have inflated the advantage observed for CICT. Three additional TAU participants withdrew before crossover to CICT. The pattern of changes after CICT in the remaining four, however, was similar to that for Immediate-CICT participants (see **Supplement 1**). In future trials, an active-comparison group appears necessary to reduce dropout.

## CONCLUSIONS

CICT has promise for reducing brain fog, improving performance of everyday tasks, and promoting return-to-work in adults with mild cognitive impairment due to PASC. These preliminary findings warrant confirmation in a large-scale RCT.

## Data Availability

All data produced in the present study are available upon reasonable request to the authors

## Acknowledgments

The conduct of this study was supported by intramural funds from the UAB Department of Psychology and UAB Integrative Center for Aging Research, and by a Scholar Award from the National Institutes of Health (NIH) National Rehabilitation Research Resource to Enhance Clinical Trials (P2CHD086951). Preparation of the manuscript was supported by grant 90IFRE0073 from the National Institute on Disability, Independent Living, and Rehabilitation Research (NIDILRR). NIDILRR is a Center within the Administration for Community Living (ACL), Department of Health and Human Services (HHS). Manuscript preparation was also supported by grant R01AG070049 from the NIH National Institute on Aging. The contents of this publication are solely the responsibility of the authors and do not necessarily represent the policy of NIDILRR, ACL, NIH, or HHS and you should not assume endorsement by the Federal Government.

## Disclosures

Posit Science donated Speed of Processing Training (SOPT) software for use in another study directed by authors GU and KB. This software was not used in this study. No restrictions on the publication or interpretation of the data from this or any other study were attached to the donation. KB serves on the Scientific Advisory Board of Posit Science, and has received travel support from Posit Science, and serves as a consultant for Posit Science. None of these activities or renumeration were for this study.

Preliminary findings from this work have been presented previously at annual meetings of the International Neuropsychological Society and American Psychological Association Division 22, but no part of the work has been published previously in a journal article or book chapter.

## Contributions

Gitendra Uswatte served as a lead for conceptualization, data curation, methodology, writing--original draft, and writing--review and editing; he contributed to data analysis, funding acquisition, project administration, and visualization. Edward Taub served as a lead for conceptualization, funding acquisition, methodology, and project administration; he contributed to writing--review and editing. Karlene Ball was a lead for conceptualization, methodology, and software; she contributed to funding acquisition, project management, and writing--review and editing. Brandon S. Mitchell and Jason A. Blake contributed to data curation, investigation, methodology, visualization, and writing--review and editing. Staci McKay was the lead for investigation and contributed to data curation, methodology, project administration, and writing--review and editing. Fedora Biney was the lead for data analysis and contributed to data curation, visualization, writing-original draft, and writing--review and editing. Olesya Iosipchuk was the lead for visualization and contributed to data analysis, data curation, writing-original draft, and writing--review and editing. Piper Hempfling contributed to investigation, writing-original, and writing--review and editing. Elise Harris contributed to data curation, writing-original, and writing--review and editing. Anne Dickerson contributed to methodology and writing--review and editing. Kristine Lokken and Amy Knight contributed to investigation, methodology, and writing--review and editing. Victor W. Mark contributed to investigation and writing--review and editing. Shruti Agnihotri contributed to investigation, writing-original draft, and writing--review and editing. Gary Cutter contributed to data analysis, visualization, and writing—review and editing.

## REFERENCES

1. Wijdicks E. Historical Lessons from Twentieth-Century Pandemics Due to Respiratory Viruses. Neurocrit Care. 2008;33(2):591–596.

2. Ford N. Long COVID and significant activity limitation among adults, by age— United States, June 1–13, 2022, to June 7–19, 2023. Morbidity and Mortality Weekly Report. 2023;72

3. Graham EL, Clark JR, Orban ZS, et al. Persistent neurologic symptoms and cognitive dysfunction in non-hospitalized Covid-19 “long haulers”. Annals of clinical and translational neurology. 2021;8(5):1073–1085.

4. Perrin PB, Ramos-Usuga D, West SJ, et al. Network Analysis of Neurobehavioral Symptom Patterns in an International Sample of Spanish-Speakers with a History of COVID-19 and Controls. International Journal of Environmental Research and Public Health. 2022;20(1):183.

5. Boldrini M, Canoll P, Klein R. How COVID-19 affects the brain. JAMA psychiatry. 2021;78(6):682–683.

6. Nouraeinejad A. Brain fog as a Long-term Sequela of COVID-19. SN Comprehensive Clinical Medicine. 2022;5(1):9.

7. Hampshire A, Trender W, Chamberlain SR, et al. Cognitive deficits in people who have recovered from COVID-19. EClinicalMedicine. 2021;39

8. Davis HE, Assaf GS, McCorkell L, et al. Characterizing long COVID in an international cohort: 7 months of symptoms and their impact. EClinicalMedicine. 2021;38

9. Jaywant A, Gunning FM, Oberlin LE, et al. Cognitive Symptoms of Post–COVID-19 Condition and Daily Functioning. JAMA Network Open. 2024;7(2):e2356098–e2356098.

10. Becker J, Lin J, Doernberg M, et al. Assessment of cognitive function in patients after COVID-19 infection. JAMA network open. 2021;4(10):e2130645–e2130645.

11. Perlis RH, Trujillo KL, Safarpour A, et al. Association of post–COVID-19 condition symptoms and employment status. JAMA network open. 2023;6(2):e2256152–e2256152.

12. Chasco E, Dukes K, Jones D, Comellas A, Hoffman R, Garg A. Brain fog and fatigue following COVID-19 infection: an exploratory study of patient experiences of long COVID. International journal of environmental research and public health. 2022;19(23):15499.

13. de Oliveira Almeida K, Nogueira Alves IG, de Queiroz RS, et al. A systematic review on physical function, activities of daily living and health-related quality of life in COVID-19 survivors. Chronic illness. 2023;19(2):279–303.

14. Mathern R, Senthil P, Vu N, Thiyagarajan T. Neurocognitive rehabilitation in COVID-19 patients: a clinical review. Southern Medical Journal. 2022;115(3):227.

15. Taub E, Uswatte G, Ball K, et al. CI Cognitive Therapy: Initial Application in a Pilot Study to Improve Cognitive Impairment in Chronic Stroke Survivors. Journal of the International Neuropsychological Society. 2023;29(1):597–598.

16. Ball K, Berch DB, Helmers KF, et al. Effects of cognitive training interventions with older adults: a randomized controlled trial. JAMA. 2002;288(18):2271–2281.

17. Ball K, Edwards J, Ross L. The Impact of Speed of Processing Training on cognitive and everyday functions. Journal of Gerontology Series B: Psychological Sciences and Social Sciences. 2007;62(Special Issue 1):19–37.

18. Taub E, Miller NE, Novack TA, et al. Technique to improve chronic motor deficit after stroke. Archives of physical medicine and rehabilitation. 1993;74(4):347–354.

19. Taub E, Uswatte G, King D, Morris D, Crago J, Chatterjee A. A placebo-controlled trial of constraint-induced movement therapy for upper extremity after stroke. Stroke. 2006;37(4):1045–1049.

20. Wolf SL, Winstein CJ, Miller JP, et al. Effect of constraint-induced movement therapy on upper extremity function 3 to 9 months after stroke: the EXCITE randomized clinical trial. JAMA. 2006;296(17):2095–2104.

21. Rebok GW, Ball K, Guey LT, et al. Active study group. Ten year effects of the advanced cognitive training for independent and vital elderly cognitive training trial on cognition and everyday functioning in older adults. Journal of the American Geriatrics Society. Journal of the American Geriatrics Society. 2014;62(1):12–24.

22. Edwards J, Wadley V, Myers R, Roenker D, Cissell G, Ball K. Transfer of a speed of processing intervention to near and far cognitive functions. Gerontology. 2002;48(5):329–340.

23. Edwards J, Wadley V, Vance D, Wood K, Roenker D, Ball K. The impact of speed of processing training on cognitive and everyday performance. Aging & Mental Health. 2005;9(3):262–271.

24. Ball K, Edwards J, Ross L, McGwin G. Cognitive training decreases motor vehicle collision involvement of older drivers. Journal of the American Geriatrics Society. 2010;58(11):2107–2113.

25. Gauthier L, Taub E, Perkins C, Ortmann M, Mark V, Uswatte G. Remodeling the brain: plastic structural brain changes produced by different motor therapies after stroke. Stroke. 2008;39(5):1520–1525.

26. Taub E, Uswatte G, Mark V, et al. Method for enhancing real-world use of a more affected arm in chronic stroke: transfer package of constraint-induced movement therapy. Stroke. 2013;44(5):1383–1388.

27. Nannoni S, de Groot R, Bell S, Markus H. Stroke in COVID-19: a systematic review and meta-analysis. International journal of stroke. 2021;16(2):137–149.

28. Jaywant A, Vanderlind W, Alexopoulos G, Fridman C, Perlis R, Gunning F. Frequency and profile of objective cognitive deficits in hospitalized patients recovering from COVID-19. Neuropsychopharmacology. 2021;46(13):2235–2240.

29. Mahon S, Faulkner J, Barker-Collo S, Krishnamurthi R, Jones K, Feigin V. Slowed information processing speed at four years poststroke: evidence and predictors from a population-based follow-up study. Journal of Stroke and Cerebrovascular Diseases. 2020;29(2):104513.

30. Nasreddine Z, Phillips N, Bédirian V, et al. The Montreal Cognitive Assessment, MoCA: a brief screening tool for mild cognitive impairment. Journal of the American Geriatrics Society. 2005;53:695–9.

31. Skinner B. The Behavior of Organisms. New York: Appleton-Century. 1938.

32. Skinner B. The Technology of Teaching. New York: Appleton-Century-Crofts. 1968.

33. Taub E, Crago JE, Burgio LD, et al. An operant approach to rehabilitation medicine: overcoming learned nonuse by shaping. Journal of the experimental analysis of behavior. 1994;61(2):281–293.

34. Carswell A, McColl M, Baptiste S, Law M, Polatajko H, Pollock N. The Canadian Occupational Performance Measure: a research and clinical literature review. Canadian journal of occupational therapy. 2004;71(4):210–222.

35. Cup EH, Scholte op Reimer WJM, Thijssen MC, van Kuyk-Minis MAH. Reliability and validity of the Canadian Occupational Performance Measure in stroke patients. Clinical rehabilitation. 2003;17(4):402–409.

36. Ohno K, Tomori K, Sawada T, Kobayashi R. Examining minimal important change of the Canadian Occupational Performance Measure for subacute rehabilitation hospital inpatients. Journal of patient-reported outcomes. 2021;5:1–10.

37. Leavitt F, Katz R. Development of the mental clutter scale. Psychological reports. 2011;109(2):445–452.

38. Michielsen H, De Vries J, Van Heck G. Psychometric qualities of a brief self-rated fatigue measure: The Fatigue Assessment Scale. Journal of psychosomatic research. 2003;54(4):345–352.

39. de Kleijn W, De Vries J, Wijnen P, Drent M. Minimal (clinically) important differences for the Fatigue Assessment Scale in sarcoidosis. Respiratory medicine. 2011;105(9):1388–1395.

40. Kroenke K, Spitzer R, Williams J, Löwe B. The patient health questionnaire somatic, anxiety, and depressive symptom scales: a systematic review. General hospital psychiatry. 2010;32(4):345–359.

41. Löwe B, Unützer J, Callahan C, Perkins A, Kroenke K. Monitoring depression treatment outcomes with the patient health questionnaire-9. Medical care. 2004;42(12):1194–1201.

42. Spitzer R, Kroenke K, Williams J, Löwe B. A brief measure for assessing generalized anxiety disorder: the GAD-7. Archives of internal medicine. 2006;166(10):1092–1097.

43. Toussaint A HP, Gumz A, Wingenfeld K, Härter M, Schramm E, Löwe B. Sensitivity to change and minimal clinically important difference of the 7-item Generalized Anxiety Disorder Questionnaire (GAD-7). Journal of Affective Disorders. 2020;265:395–401.

44. Costa S, Genova H, DeLuca J, Chiaravalloti N. Information processing speed in multiple sclerosis: Past, present, and future. Multiple Sclerosis Journal. 2017;23(6):772–789.

45. Jaywant A, Barredo J, Ahern D, Resnik L. Neuropsychological assessment without upper limb involvement: a systematic review of oral versions of the Trail Making Test and Symbol-Digit Modalities Test. Neuropsychological rehabilitation. 2018;28(7):1055–1077.

46. Benedict R, DeLuca J, Phillips G, et al. Validity of the Symbol Digit Modalities Test as a cognition performance outcome measure for multiple sclerosis. Multiple Sclerosis Journal. 2017;23(5):721–733.

47. Krishnan K, Rossetti H, Hynan L, et al. Changes in Montreal Cognitive Assessment scores over time. Assessment. 2017;24(6):772–777.

48. Cohen J. Statistical power analysis for the behavioral sciences. 2nd ed. Lawrence Erlbaum Associates; 1988.

49. Chandler J, Cumpston M, Li T, Page M, Welch V. Cochrane handbook for systematic reviews of interventions. Wiley; 2019.

50. Uswatte G, Taub E, Mark V, Perkins C, Gauthier L. Central nervous system plasticity and rehabilitation. 2 ed. Handbook of rehabilitation psychology. American Psychological Association; 2010.

51. Ungerleider L, Doyon J, Karni A. Imaging brain plasticity during motor skill learning. Neurobiology of learning and memory. 2002;78(3):553–564.

52. Kleim J, Barbay S, Nudo R. Functional reorganization of the rat motor cortex following motor skill learning. Journal of neurophysiology. 1998;80(6):3321–3325.

