## Supplement 1 for "Long COVID Brain Fog Treatment: Findings from a Pilot Randomized Controlled Trial of Constraint-Induced Cognitive Therapy"

**METHODS**

**Study Design**
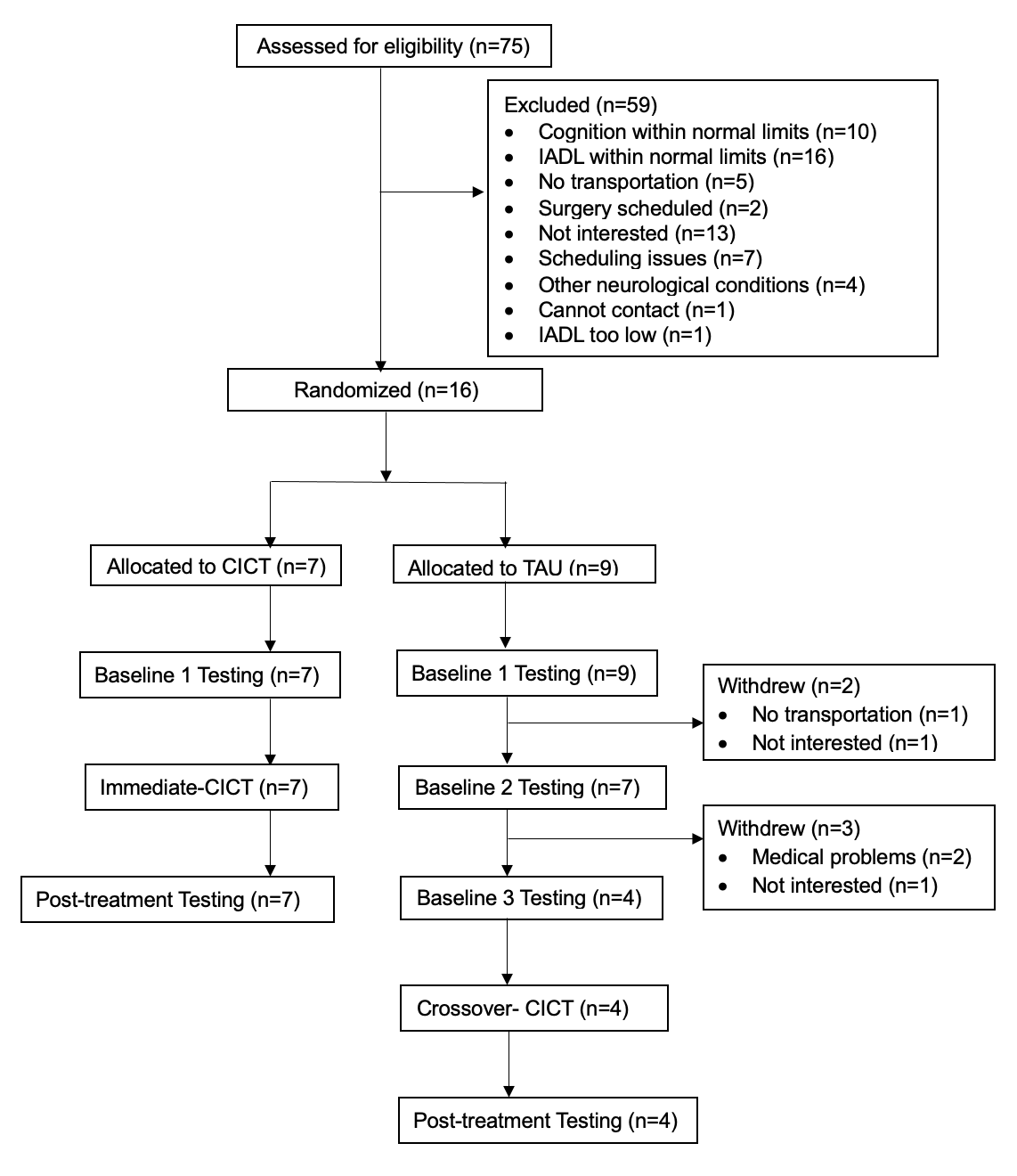

**Figure s1. Study Flowchart.** CICT indicates Constraint-Induced Cognitive Therapy; IADL, instrumental activities of daily living; TAU, treatment-as-usual.

| Participant | Baseline 1 | Post-treatment | Change Score | Effect Size (*d'*)^a^ | Improvement ≥ MCID |
| --- | --- | --- | --- | --- | --- |
| Immediate-CICT group | | | | | |
| S1 | 23 | 21 | -2 |  | No |
| S2 | 25 | 30 | 5 |  | Yes |
| S3 | 25 | 27 | 2 |  | Yes |
| S4 | 23 | 21 | -2 |  | No |
| S5 | 25 | 28 | 3 |  | Yes |
| S6 | 20 | 29 | 9 |  | Yes |
| S7 | 24 | 27 | 3 |  | Yes |
| Summary^b^ | 23.6 (1.8) | 26.1 (3.7) | 2.6 (3.9) | 0.7 | 71% |
| TAU group | | | | | |
| S8^c^ | 16 | 25 | 9 |  | Yes |
| S9 | 26 | 26 | 0 |  | No |
| S10 | 22 | 25 | 3 |  | Yes |
| S11 | 25 | 26 | 1 |  | No |
| S12 | 25 | 24 | -1 |  | No |
| S13 | 26 | 26 | 0 |  | No |
| S14 | 26 | 29 | 3 |  | Yes |
| Summary^b^ | 25 (1.6) | 26 (1.7) | 1 (1.7) | 0.6 | 33% |
| Abbreviations: CICT, Constraint-Induced Cognitive Therapy; MCID, minimal clinically important difference; TAU, treatment-as-usual.  ^a^ *d'* is a repeated-measures effect size index; it is the mean change divided by its standard deviation. Values ≥ 0.57 are considered large.  ^b^ Values are *M* (*SD*) unless otherwise indicated.  ^c^ S8 was excluded from the calculation of the summary variables because their Baseline 1 MoCA score was an outlier. S8’s score fell below the lower bound of the Inter-quartile Range minus 1.5 times the Inter-quartile Range.^1^ Another perspective is that all the other participants had scores in the mild cognitive impairment range (18-26), while S8 had a score in the moderate range (10-17).^2^ The summary variable values for the TAU group with S8 included were [*M* (*SD*)]: Baseline 1, 23.7 (3.7); Post-treatment, 25.9 (1.6); change score, 2.1 (3.4); *d'* = 0.6. | | | | | |

**Participants**

**Table s1. Montreal Cognitive Assessment (MoCA) Scores for Immediate-CICT and**

**TAU Participants**

**Intervention: Constraint-Induced Cognitive Therapy (CICT)**

Speed of Processing Training

SOPT involves presenting a visual target on a computer screen. The program consists of three conditions that increase in cognitive load.^3^ In the first task, Stimulus Detection, participants are asked to identify a central target at an increasingly faster rate. Once participants master the task at the shortest stimulus duration, they move to the second task, Divided Attention. At this stage, they are asked to divide their attention between detecting a central target and localizing a peripheral target. Once this task is mastered, they move to the third and final task – Selective Attention. This task is considered the hardest as it adds visual distractors of varying conspicuity to the stimulus. Each of the three tasks get progressively more difficult, including importantly by presenting stimuli more rapidly, as participants provide correct answers.

Training by In-Lab Shaping of Instrumental Activities of Daily Living (IADL)

On the first day of training, participants are asked to identify everyday cognition-based tasks are challenging or beyond their current abilities. During subsequent training sessions, trainers and participants work together to improve the performance of the identified tasks. The training is based on shaping principles. Small improvements are rewarded by providing praise and warmly encouraging participants to improve their performance. Errors are not commented on (i.e., punished). Performance time is recorded and shared with the participant after each trial. Participants are encouraged to improve their speed of task completion, while verbal feedback ensures that the participant maintains the quality of their performance. The tasks used come from a bank of standard cognitive tasks for which formal shaping plans have been formulated. New shaping tasks can be created to accommodate the participant’s needs and preferences.

Transfer Package

The TP bridges the gap between the in-lab and the real-world performance. More specifically, it aims to transfer therapeutic gains achieved in the lab to real-life situations through the use of behavioral techniques to reinforce treatment adherence. **Table s2** describes individual elements of the TP. The TP was initially developed to help participants with upper-extremity hemiparesis due to stroke transfer gains from in-lab physical training to everyday life.^4^ The TP procedures, such as behavioral contract, are common in behavioral therapies that target substance abuse and medication adherence.^5-7^

**Supplement 1** continues on next page.

**Table s2. Elements of the Cognitive Transfer Package**

| **Transfer Package Element** | **Description** |
| --- | --- |
| Behavioral Contract | At the outset of treatment, trainers negotiate a “contract” with participants and their family members or other informal caregivers about their responsibilities, as well as the responsibilities of the treatment team. This written agreement specifies activities with important cognitive components that are important to participants and commits participants to attempt these activities outside the treatment setting and to attend and engage in all training sessions. The agreement, in addition, specifies when support from a family member is needed to carry out an activity safely. |
| CTAL and INCA | The CTAL and INCA, which are completed at the beginning of each training session, collect information about the performance of everyday activities with important cognitive components outside the treatment setting by participants (see **Feasibility** section of this supplement). These structured interviews serve as occasions to (a) reward participants with verbal praise for attempts to carry out cognition-based activities in daily life and (b) support problem solving by participants and their family caregivers, when relevant, regarding perceived and actual barriers to doing so. |
| Home Skill Assignment | At the end of each training session, participants, with support from family caregivers, if needed, are asked to carry out up to 10 cognition-based tasks at home before the next session. The tasks are selected from a bank developed by this lab; the list is refreshed, as needed, at each session. Participants check off the activities carried out on the form provided to them, plus enter when the activities were carried out and comment on any difficulties encountered. Examples of tasks are: prepare a meal, sort mail, write emails, play board games, count calories, make a grocery list, make a budget for the week, pay a bill, and organize closet. Completion of the activities assigned is reviewed at the outset of each training session. |
| Problem Solving | Throughout each session, but particularly during the CTAL and INCA interviews and review of home skill assignments, trainers support problem solving by participants regarding any perceived or actual barriers to the performance of cognition-based tasks in daily life. The problem-solving procedures are based on the “goal-plan-do-check” procedure employed by Skidmore and colleagues.^8^ |
| Family Engagement | Trainers coach family caregivers to (a) permit clients to carry out tasks independently when safe to do so, (b) provide clients with supervised practice in the home, and (c) prompt clients to perform IADL at home. |
| Follow-up Program | Trainers give participants an individualized program of 10 to 15 tasks to practice for a total of 30-60 minutes a day at home at the end of the last training session. Follow-up telephone calls are scheduled weekly for the first month after treatment. During each call, trainers conduct the CTAL and INCA interviews, review and encourage compliance with participants’ home programs, and modify the programs as needed, e.g., replacing easy with difficult tasks. As in training sessions, the CTAL and INCA interviews are occasions to reward participants with verbal praise for engaging in cognition-based activities and to support problem solving by participants regarding any barriers to doing so. |

Abbreviations: CTAL, Cognitive Task Activity log; INCA, Inventory of New Cognitive Activities; IADL. Instrumental Activities of Daily Living

**Typical CICT Training Session**

INCA and CTAL administration, review of homework, discussion of compliance with behavioral contract (45 min)

Rest (15 min)

SOPT (45 min)

Rest (15 min)

Training by shaping of everyday cognitive tasks (45 min)

Review of the day, discussion of that day's homework assignment, problem solving (15 min)

***Figure s2***. **Typical Constraint Induced Cognitive Therapy Session Schedule.** CTAL indicates Cognitive Task Activity Log; INCA, Inventory of New Cognitive Activities; SOPT, Speed of Processing Training.

**Feasibility**

The INCA and CTAL were used to track engagement by participants in CICT. The POS was used to assess participants’ acceptance of CICT. Each is described below.

Inventory of New Cognitive Activities

The INCA was developed by this lab to track resumption of or improvement in a wide range of functional, cognition-based, activities outside the treatment setting. In this structured interview, participants report (a) resumption of any activities not performed since stroke onset, i.e., new activities, and (b) meaningful improvement in any activities since their last INCA interview. To verify their reports, participants are asked to demonstrate or explain how the activities are done. The number of new activities is counted on each interview occasion and added to the cumulative total from the previous occasion; the same is done for improved activities. An example of a completed INCA is provided in **Figure s3**.

**Supplement 1** continues on next page.

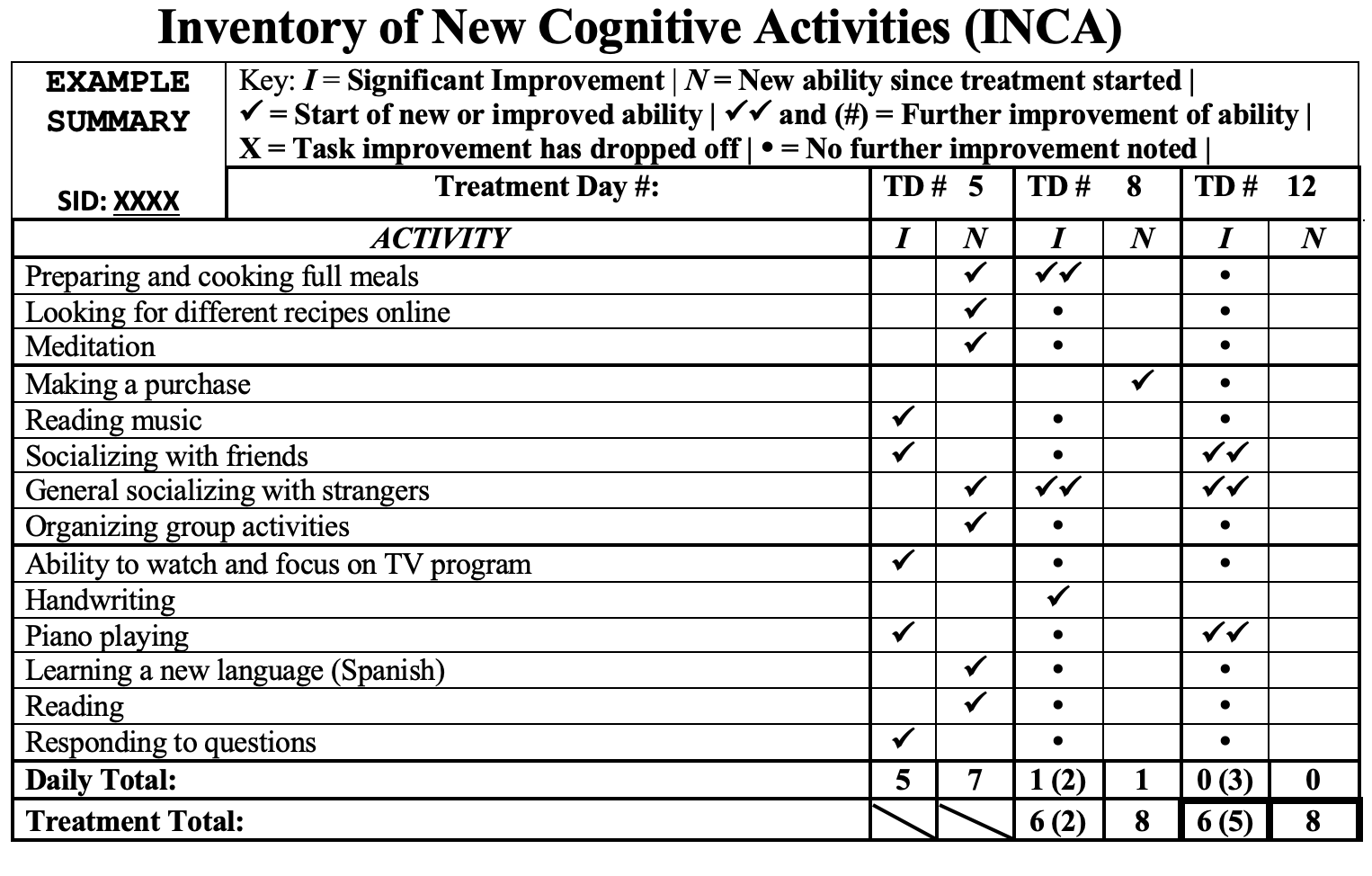

**Figure s3**. **Example of completed INCA data collection form.**

Cognitive Task Activity Log

The CTAL is a structured interview that was modeled after the Motor Activity Log (MAL)^9,10^ and the Verbal Activity Log (VAL),^11^ both of which have rigorous evidence of validity.^9-11^ Participants are asked to rate their degree of independence (Independence Scale) and quality of their performance (Quality Scale) on 24 cognition-based activities. The questionnaire is scored on a five-point scale (0=activity not done at all, 5=completed activity normally). The total score is the average of the Independence and Quality scores. **Tables s3** and **s4** show individual items and scales for CTAL, respectively.

| Item | Description |
| --- | --- |
| 1 | Start a conversation with a person outside the home |
| 2 | Remember the day of the week |
| 3 | Organize medications to take |
| 4 | Pay attention to a task with several steps |
| 5 | Use a smartphone or computer to assess multiple websites |
| 6 | Make a purchase using cash or a credit card |
| 7 | Remember appointments or events |
| 8 | Detailed and correct responses to questions |
| 9 | Look up a phone number or cell phone contact |
| 10 | Calculate a tip in a restaurant |
| 11 | Manage day-to-day purchases without overspending or forgetting (e.g., groceries) |
| 12 | Remembering personal effects (e.g., keys, wallet, purse) |
| 13 | Navigate to a location beyond walking distance |
| 14 | Remember to take meds according to directions |
| 15 | Follow or understand the plot of a movie |
| 16 | Use a keypad (e.g., remote, microwave) |
| 17 | Prepare food that includes at least three ingredients |
| 18 | Read and understand written text (e.g., magazine, book, newspaper) |
| 19 | Writing or typing multi-word messages (e.g., email, letter, note) |
| 20 | Remember PIN number (e.g., SmartPhone, debit card, security code) |
| 21 | Remember passwords for multiple websites |
| 22 | Locate an item on a file system, physical or computer |
| 23 | Check accuracy of an account balance or billing statement (e.g., checking, saving, Greenphire) |
| 24 | Put away items (e.g., clothes, linen) |

**Table s3. Cognitive Task Activity Log (CTAL) Items**

**Table s4. Cognitive Task Activity Log (CTAL) Quality and Independence scales**

| Score | Description |
| --- | --- |
| CTAL Quality Scale | |
| 0 | Activity not done at all (never) |
| 1 | Tried to do the activity, but was unable to complete it (very poor) |
| 2 | Sometimes completed the activity, but it was very slow or difficult (poor) |
| 3 | Routinely completed the whole activity, but it was slow or moderately difficult (fair) |
| 4 | Always completed the activity, but not as rapidly or easily as normal (almost normal) |
| 5 | Always completed the activity as well and as easily as normal (normal) |
| CTAL Independence Scale | |
| A. Prompt: Activity required prompting or reminder to start… | |
| 0 | All the time |
| 1 | Almost all the time |
| 2 | Most of the time |
| 3 | About half the time |
| 4 | Less than half the time |
| 5 | None of the time |
| B. Prompt: Activity requires assistance or supervision to complete… | |
| 0 | All the time |
| 1 | Almost all the time |
| 2 | Most of the time |
| 3 | About half the time |
| 4 | Less than half the time |
| 5 | None of the time |
| Note. Codes for recording “no” responses: (1) “I never do that activity, with or without help, because it is not relevant.” For example, a person does not have or use a smartphone or a computer. (assign N/A and drop the item in future administrations), (2) “Someone else did the activity for me.” (assign a “0”), and (3) “I sometimes do that activity but did not have the opportunity since the last time I answered these questions.” (carry-over last assigned number for that activity.)  If participants debate between two scores, they are allowed to choose a score in between (e.g., 3.5).  The CTAL Quality test score is the average of the 24 item scores.    The CTAL Independence scale is divided into two parts, A and B, that consist of two different questions. The CTAL Independence test score is calculated by taking the minimum of the question A and B scores for each item and then averaging these values. The CTAL Total test score is the average of the Independence and Quality test scores. | |

Participant Opinion Survey

The POS is a survey developed by this lab to assess participants' satisfaction with the treatment program. POS asks caregivers and patients to rate the treatment’s difficulty, benefit, and satisfaction levels on a scale from 1 (not at all) to 7 (extremely) before and after the treatment (see **Table s5**).

**Table s5. Participant Opinion Survey Items**

| Item | Description |
| --- | --- |
| Participant Form | |
| 1 | How difficult do you think your therapy program has been? |
| 2 | I believe that the therapy program has benefitted me. |
| 3 | How satisfied are you with your therapy program? |
| Caregiver Form | |
| 1 | I believe that the therapy program has benefitted the participant. |
| Note. For questions 1 and 3 in the participant form, a 7-point scale was used where 1 was “not at all” and 7 was “extremely”. For question 2 in the participant form and question 1 in the caregiver form, response anchors were “strongly disagree” and “strongly agree” for 1 and 7, respectively. | |

**Data Analysis**

TAU Crossover to CICT

TAU group completed three baseline assessments prior to crossover to CICT; a fourth assessment was completed after CICT (See Figure s1). The third baseline assessment was chosen for comparison against post-treatment scores on all outcome measures as this accounted best for the cumulative practice effects of all baseline assessments. The small sample size (*n* = 4) precluded formal inferential statistical analysis. Instead, several descriptive statistics were calculated, i.e., the mean change from Baseline 3 to Post-treatment along with the corresponding *SD* and effect size index (*d')*. (*d'* is the mean change divided by its SD; values ≥0.57 are considered large.)^12^ In addition, the number of participants with improvement greater than a minimal clinically important difference (MCID) was counted. Last, spaghetti plots were drawn.

**RESULTS**

**Feasibility**

Engagement by Participants in CICT

Participants were highly engaged in CICT. According to the INCA, Immediate-CICT participants resumed, on average, 14 cognition-based activities (*SD* = 6.7, range = 7-26; **Figure s4**) after starting therapy that participants had ceased post-COVID; the corresponding value for Crossover-CICT participants was 8 (*SD* = 2.9, range = 5-11; **Figure s5)**. In addition, according to the INCA, Immediate-CICT participants improved their performance of 85 cognition-based activities (*SD* = 72.9, range = 17-192; **Figure s6**) after starting therapy; the corresponding value for Crossover-CICT participants was 23 (*SD* = 25.8, range = 6-61; **Figure s7**). As noted, the CTAL measures how well and independently participants perform 24 cognition-based activities outside of the treatment setting; the Total score ranges from 0 (activity not done at all) to 5 (activity completed normally). At the beginning of treatment, the mean Total in the Immediate-CICT group was 3.2 (*SD* = 0.5, range = 2.4-3.9); at the end of treatment the mean Total in this group was 4.4 (*SD* = 0.5, range 3.8-4.8), which is close to normal (**Figure s8**). Corresponding values at the beginning and end of treatment in the Crossover-CICT group were 2.9 (*SD* = 1.1, range = 1.3-3.6) and 4 (*SD* = 0.7, range = 3.0-4.5), respectively (**Figure s9**). In both the Immediate- and Crossover-CICT groups, a ceiling effect restricted the observation of improvement on the CTAL; about half the participants in each group achieved scores above 4 midway through therapy.

New Activities

| Training Hours | S1 | S2 | S3 | S4 | S5 | S6 | S7 | #REF! | #REF! | #REF! | #REF! | Mean |
| --- | --- | --- | --- | --- | --- | --- | --- | --- | --- | --- | --- | --- |
| 5 | 4 | 0 | 3 | 5 | 1 | 1 | 1 | 1 | 1 | 1 | 1 | 2.14286 |
| 10 | 7 | 4 | 5 | 8 | 1 | 5 | 3 |  |  |  |  | 4.71429 |
| 15 | 10 | 5 | 11 | 8 | 6 | 7 | 5 |  |  |  |  | 7.42857 |
| 20 | 15 | 6 | 12 | 10 | 7 | 7 | 5 |  |  |  |  | 8.85714 |
| 25 | 15 | 7 | 12 | 10 | 9 | 7 | 7 |  |  |  |  | 9.57143 |
| 30 | 18 | 14 | 12 | 10 | 15 | 7 | 7 |  |  |  |  | 11.8571 |
| 35 | 26 | 17 | 16 | 10 | 15 | 7 | 7 |  |  |  |  | 14 |

**Fig s4. Everyday Cognition-based Activities Resumed During Immediate-CICT.** The number of activities resumed after starting therapy that participants had ceased post-COVID was assessed with the Inventory of New Cognitive Activities.

New Activities

| Training Hours | #REF! | #REF! | #REF! | #REF! | #REF! | #REF! | #REF! | S9 | S10 | S11 | S13 | Mean |
| --- | --- | --- | --- | --- | --- | --- | --- | --- | --- | --- | --- | --- |
| 5 | 1 | 1 | 1 | 1 | 1 | 1 | 1 | 2 | 2 | 4 | 4 | 3 |
| 10 |  |  |  |  |  |  |  | 4 | 2 | 5 | 5 | 4 |
| 15 |  |  |  |  |  |  |  | 8 | 5 | 5 | 5 | 5.75 |
| 20 |  |  |  |  |  |  |  | 9 | 5 | 5 | 6 | 6.25 |
| 25 |  |  |  |  |  |  |  | 9 | 9 | 5 | 6 | 7.25 |
| 30 |  |  |  |  |  |  |  | 9 | 9 | 5 | 6 | 7.25 |
| 35 |  |  |  |  |  |  |  | 10 | 11 | 5 | 6 | 8 |

**Fig s5. Everyday Cognition-based Activities Resumed During Crossover-CICT.** The number of activities resumed after starting therapy that participants had ceased post-COVID was assessed with the Inventory of New Cognitive Activities.

Improved Activities

| Training Hours | S1 | S2 | S3 | S4 | S5 | S6 | S7 | #REF! | #REF! | #REF! | #REF! | Mean |
| --- | --- | --- | --- | --- | --- | --- | --- | --- | --- | --- | --- | --- |
| 5 | 0 | 3 | 2 | 1 | 2 | 0 | 0 | 1 | 1 | 1 | 1 | 1.14286 |
| 10 | 11 | 16 | 6 | 11 | 5 | 6 | 7 |  |  |  |  | 8.85714 |
| 15 | 29 | 29 | 17 | 20 | 23 | 14 | 10 |  |  |  |  | 20.2857 |
| 20 | 61 | 45 | 31 | 28 | 28 | 14 | 15 |  |  |  |  | 31.7143 |
| 25 | 75 | 63 | 44 | 30 | 37 | 17 | 26 |  |  |  |  | 41.7143 |
| 30 | 127 | 123 | 55 | 30 | 50 | 17 | 30 |  |  |  |  | 61.7143 |
| 35 | 192 | 180 | 91 | 30 | 50 | 17 | 35 |  |  |  |  | 85 |

**Fig s6. Everyday Cognition-based Activities Improved During Immediate-CICT.** The number of activities improved after starting therapy was assessed with the Inventory of New Cognitive Activities. The lines for S1 and S2 are cut off because they extend beyond the range that it is possible to graph without obscuring the others’ data. The number of improved activities at the end of therapy for S1 and S2 were 192 and 180, respectively.

Improved Activities

| Training Hours | #REF! | #REF! | #REF! | #REF! | #REF! | #REF! | #REF! | S9 | S10 | S11 | S13 | Mean |
| --- | --- | --- | --- | --- | --- | --- | --- | --- | --- | --- | --- | --- |
| 5 | 1 | 1 | 1 | 1 | 1 | 1 | 1 | 0 | 1 | 5 | 0 | 1.5 |
| 10 |  |  |  |  |  |  |  | 3 | 2 | 12 | 0 | 4.25 |
| 15 |  |  |  |  |  |  |  | 4 | 13 | 15 | 2 | 8.5 |
| 20 |  |  |  |  |  |  |  | 7 | 20 | 16 | 3 | 11.5 |
| 25 |  |  |  |  |  |  |  | 8 | 37 | 16 | 6 | 16.75 |
| 30 |  |  |  |  |  |  |  | 8 | 44 | 17 | 6 | 18.75 |
| 35 |  |  |  |  |  |  |  | 8 | 61 | 17 | 6 | 23 |

**Fig s7. Everyday Cognition-based Activities Improved During Crossover-CICT.** The number of activities improved after starting therapy was assessed with the Inventory of New Cognitive Activities.

CTAL Total scale, points

| Training Hours | S1 | S2 | S3 | S4 | S5 | S6 | S7 | Mean |
| --- | --- | --- | --- | --- | --- | --- | --- | --- |
| 0 | 3.9 | 3.22 | 3.1 | 3.2 | 2.4 | 3.1 | 3.6 | 3.21714 |
| 5 | 3.17 | 3.455 | 3.9 | 3.23 | 2.825 | 2.195 | 4.02 | 3.25643 |
| 10 | 3.3 | 3.435 | 4.46 | 3.8 | 3.24 | 3.18 | 4.155 | 3.65286 |
| 15 | 3.555 | 3.77 | 4.77 | 4.26 | 4.32 | 3.3 | 4.25 | 4.03214 |
| 20 | 3.645 | 3.795 | 4.865 | 3.96 | 4.63 | 3.55 | 4.47 | 4.13071 |
| 25 | 4.08 | 4.15 | 4.96 | 4.18 | 4.6 | 3.72 | 4.55 | 4.32 |
| 30 | 4.12 | 4.38 | 4.96 | 4.24 | 4.78 | 4.03 | 4.64 | 4.45 |
| 35 | 4.455 | 4.54 | 4.96 | 4.3 | 4.78 | 4.03 | 4.64 | 4.52929 |

**Fig s8. Engagement in IADL Outside the Treatment Setting by Immediate-CICT Group.** The Cognitive Task Activity Log (CTAL) Total score indexes how independently and well respondents carry out 24 cognition-based activities outside the treatment setting. A 0 indicates that an activity was not done at all; a 5 indicates that the activity was completed normally. A ceiling effect restricted the observation of improvement for several participants; three of the seven had test scores above 4 midway through therapy.

CTAL Total scale, points

| Training Hours | S9 | S10 | S11 | S13 | Mean |
| --- | --- | --- | --- | --- | --- |
| 0 | 3.5 | 3.3 | 3.6 | 1.25 | 2.9125 |
| 5 | 3.805 | 2.955 | 3.45 | 1.06 | 2.8175 |
| 10 | 3.93 | 2.83 | 4.465 | 1.885 | 3.2775 |
| 15 | 4.24 | 3.575 | 4.595 | 2.145 | 3.63875 |
| 20 | 4.365 | 3.19 | 4.72 | 2.48 | 3.68875 |
| 25 | 4.59 | 3.55 | 4.83 | 2.67667 | 3.91167 |
| 30 | 4.66 | 3.68 | 4.91 | 2.67667 | 3.98167 |
| 35 | 4.63 | 3.805 | 4.91 | 2.67667 | 4.00542 |

**Fig s9. Engagement in IADL Outside the Treatment Setting by Crossover-CICT Group.** The Cognitive Task Activity Log (CTAL) Total score indexes how independently and well respondents carry out 24 cognition-based activities outside the treatment setting. A 0 indicates that an activity was not done at all; a 5 indicates that the activity was completed normally. A ceiling effect restricted the observation of improvement for half of the participants; two of four had test scores above 4 midway through therapy.

**Efficacy: Everyday Activities**

COPM Satisfaction Scale

There was a main effect of group assignment on post-treatment scores, *F*(1,9) = 22.8, *p* < 0.001; Immediate-CICT participants, compared to TAU participants, reported a very large improvement in satisfaction with their performance of everyday cognition-based activities: *MD* = 4.0 points; *95% CI*, 2.4 to 5.6; *d* = 2.6. In addition, there was a significant interaction between group assignment and baseline COPM Satisfaction scores, *F*(1, 9) = 6, *p* = 0.037; the advantage of CICT over TAU was larger for participants with low baseline scores than for those with high baseline scores (see **Figure s10**). The results reported above were from an ANOVA model using raw scores; results from a parallel ANOVA model substituting ranks for raw scores were similar, *F*(1, 9) = 5.64 *p* = 0.04. An analysis of variance (ANOVA) model with an interaction term (Group x Baseline 1) rather than an analysis of covariance (ANCOVA) model was employed here because homogeneity of variance was not present.

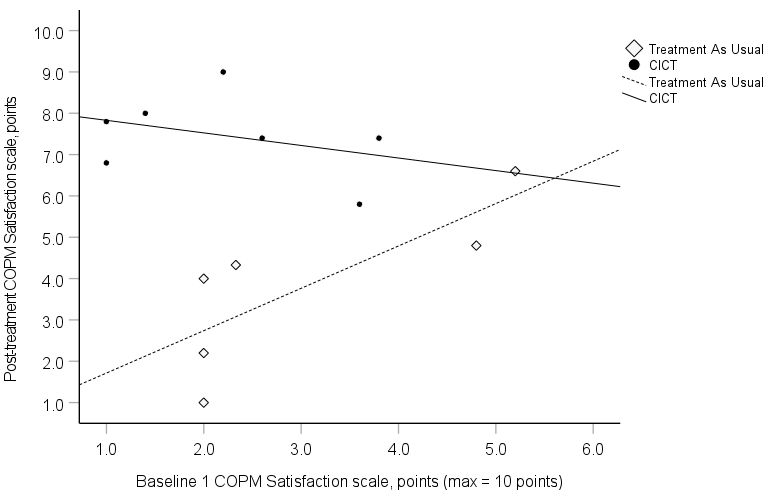

**Figure s10. Satisfaction with Performance of Everyday Cognition-based Activities**. The Canadian Occupational Performance Measure (COPM) Satisfaction scale measures how satisfied participants are with the performance of 5 self-selected, cognition-based activities. All of the Immediate-Constraint-Induced Cognitive Therapy (CICT) participants had clinically meaningful improvements; only two of six Treatment-As-Usual (TAU) participants with COPM data did.

**Efficacy: Psychological Distress**

Fatigue Symptom Severity Before and After CICT and TAU

There was a main effect of group assignment on post-treatment Fatigue Assessment Scale (FAS) scores, *F*(1,10) = 7.6, *p* = 0.02; Immediate-CICT participants, compared to TAU participants, reported very large reductions in fatigue symptoms: *MD* = -10.9 points; *95% CI*, -15.6 to -6.3; *d*= -1.8. There was also a significant interaction between group assignment and baseline FAS scores, *F*(1, 10) = 11.9, *p* = 0.006; the advantage of Immediate-CICT over TAU was larger for participants with high baseline scores than for those with lower baseline scores (see **Figure s11**).

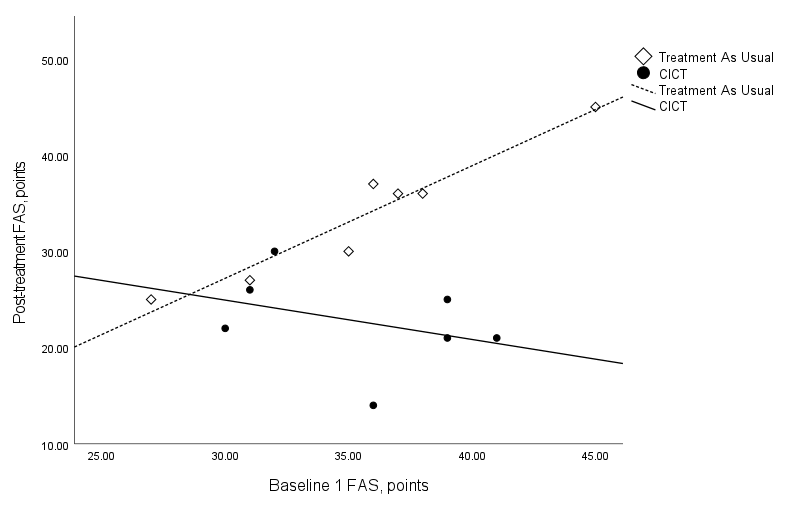

**Figure s11. Fatigue Symptom Severity.** The Fatigue Assessment Scale (FAS) was used to assess fatigue symptom severity. On the FAS, respondents are asked to rate how frequently 10 symptoms are experienced using a 5-point scale (1 = never, 5 = always). The test score is the sum of the item scores. Six of seven Immediate-Constraint-Induced Cognitive Therapy (CICT) participants had clinically meaningful improvements; only two of seven Treatment-As-Usual (TAU) participants did.

Depressive and Anxiety Symptom Severity Before and After CICT and TAU: Bar Graphs

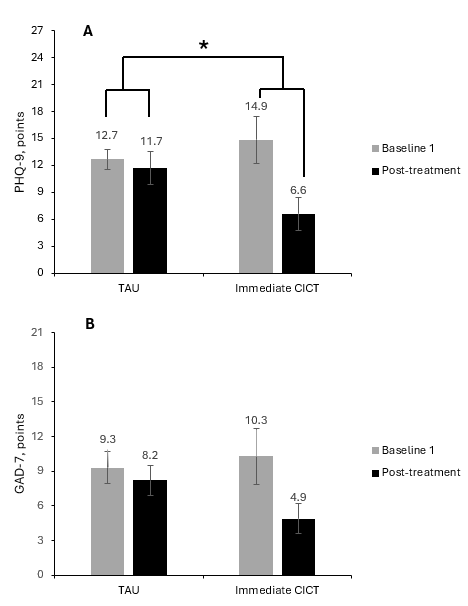

**Figure s12. (A) Depressive and (B) Anxiety Symptom Severity Before and After Constraint-Induced Cognitive Therapy (CICT) and Treatment-As-Usual (TAU).** The Patient Health Questionnaire-9 (PHQ-9) and General Anxiety Disorder-7 (GAD-7) were used to assess depression and anxiety symptom frequency, respectively. The PHQ-9 lists nine symptoms; the GAD-7 lists seven. On both surveys, respondents rate how frequently the symptoms are experienced using 4-point scales (0, never; 3, nearly every day). The test score is the sum of the item-scores. Panel A plots raw scores as opposed to ranks (see next section). Four of seven Immediate-CICT participants had clinically meaningful improvements in depressive symptoms; only two of seven TAU participants did. Five of seven Immediate-CICT participants had clinically meaningful improvements in anxiety symptoms; only two of seven Treatment-As-Usual (TAU) participants did

**p* < .05

Results of Alternate Analysis of the Patient Health Questionnaire-9 (PHQ-9) Data

As noted in the **Data Analysis**, the PHQ-9 data were not distributed normally, which violated a requirement for standard analysis of covariance (ANCOVA). Hence, we conducted an ANCOVA test substituting ranks for raw scores. The latter detected a statistically significant advantage for Immediate-CICT over TAU; the test statistic and *p* value from this test are reported in the **Results**, along with the median post-treatment PHQ-9 score and corresponding inter-quartile range (IQR) in each group. At baseline, the median PHQ-9 score in the Immediate-CICT group was 15 (IQR = 8 – 21); the corresponding value in the TAU group was 12 (IQR = 10 – 15). To be transparent, we here report the standard ANCOVA test results, which were negative: *MD* = -5.1; *95%* *CI*, -10.7 - 0.4; *F*(1,11) =.2, *p* = 0.07; *d* = -0.9.

**Efficacy: In-lab Cognitive Testing**

Results of Alternate Analysis of the Symbol Digit Modalities Test (SMDT) Data

As noted in the **Data Analysis**, the SDMT data were not distributed normally, which violated a requirement for standard analysis of covariance (ANCOVA). Hence, we conducted an ANCOVA test substituting ranks for raw scores. The latter detected a statistically significant advantage for Immediate-CICT over TAU; the test statistic and *p* value from this test are reported in the **Results**, along with the median post-treatment SMDT score and corresponding IQR in each group. At baseline, the median SDMT score in the Immediate-CICT group was 47 (IQR = 30 – 49); the corresponding value in the TAU group was 43 (IQR = 36 – 50). To be transparent, we here report the standard ANCOVA test results, which were negative: SDMT *MD* = 7.8; *95% CI*, -2.1 to 17.8; *F*(1,10) = 3.1, *p* = 0.11; *d*=0.6 (see **Figure s13)**.

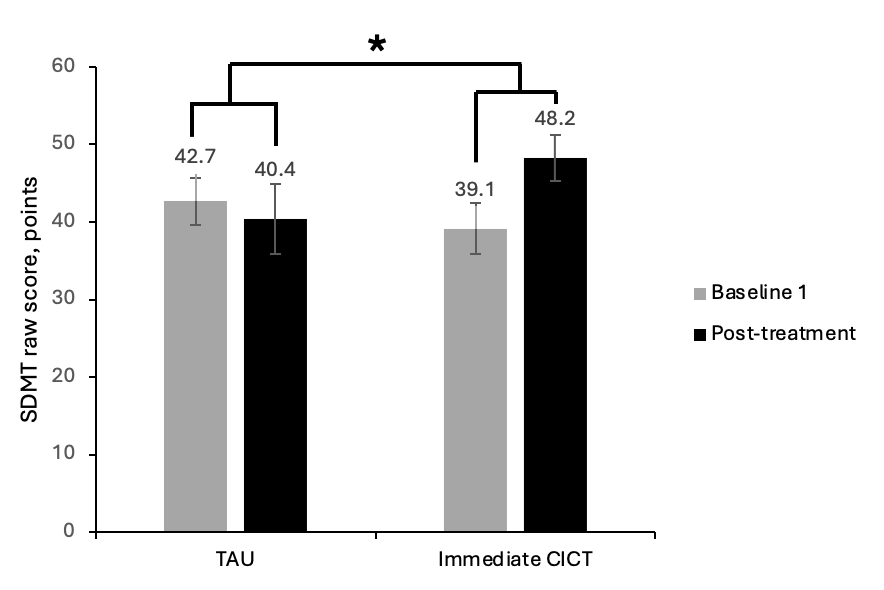

**Figure s13**. **Cognitive Processing Speed Before and After Immediate-Constraint-Induced Cognitive Therapy (CICT) and Treatment-As-Usual (TAU).** The Symbol Digit Modalities Test (SDMT) was used to assess cognitive processing speed. The total score is the number of symbols that are coded correctly in 90 s (max = 110). Raw as opposed to rank scores are plotted. Four of seven Immediate-CICT participants had clinically meaningful improvements in cognitive processing speed, while three of six TAU participants did.

**p* < .05

**Crossover CICT Results**

As noted, four TAU participants completed Baseline 3 testing and received CICT. **Table s6** shows mean pre- to post-CICT changes and corresponding effect sizes for these participants**. Figures s14-s21** display participant-level data for each outcome measure. **Table s7** shows the employment status of the TAU group before and after COVID-19 onset and after CICT.

**Table s6. Mean (*SD*) Test Scores of TAU Participants Before and After Crossover to CICT**

| Outcome | Baseline 3 | Post-treatment | Change | Effect size (*d'*) | Improvement ≥ MCID^a^ |
| --- | --- | --- | --- | --- | --- |
| COPM Performance scale, points, range 1-10 | 4.6 (2.3) | 6.4 (2.2) | 1.9 (1.2) | 1.5 | 75% |
| COPM Satisfaction scale, points, range 1-10 | 3.1 (1.6) | 5.9 (2.4) | 2.8 (1.0) | 2.8 | 75% |
| MCS, points, range 1-10 | 6.9 (1.6) | 4.1 (1.1) | -2.8 (2.4) | -1.2 | N/A |
| FAS, points, range 1-50 | 30 (11.2) | 26 (11.5) | -4 (4.8) | -0.8 | 25% |
| PHQ-9, points, 0-27 | 11.8 (7.6) | 8 (7.4) | -3.8 (4.3) | -0.9 | 25% |
| GAD-7, points, range 0-21 | 9.0 (5.4) | 7.0 (4.1) | -2.0 (3.4) | -0.6 | 50% |
| SDMT raw scores, points, range 0-110 | 46 (14.8) | 50.3 (9.5) | 4.3 (8.5) | 0.5 | 25% |
| MoCA, points, range 0-30 | 25.3 (2.6) | 27.8 (2.1) | 2.5 (1.3) | 1.9 | 75% |
| Abbreviations: COPM, Canadian Occupational Performance Measure; MCS, Mental Clutter Scale; FAS, Fatigue Assessment Scale; PHQ-9, Patient Health Questionnaire-9; GAD-7, Generalized Anxiety Disorder-7; SDMT, Symbol Digit Modalities Test; MoCA, Montreal Cognitive Assessment.  ^a^ This is the number of participants with improvement above the threshold for an MCID expressed as a percentage of the number of participants for whom data is available. No validated MCID is available for the MCS. | | | | | |

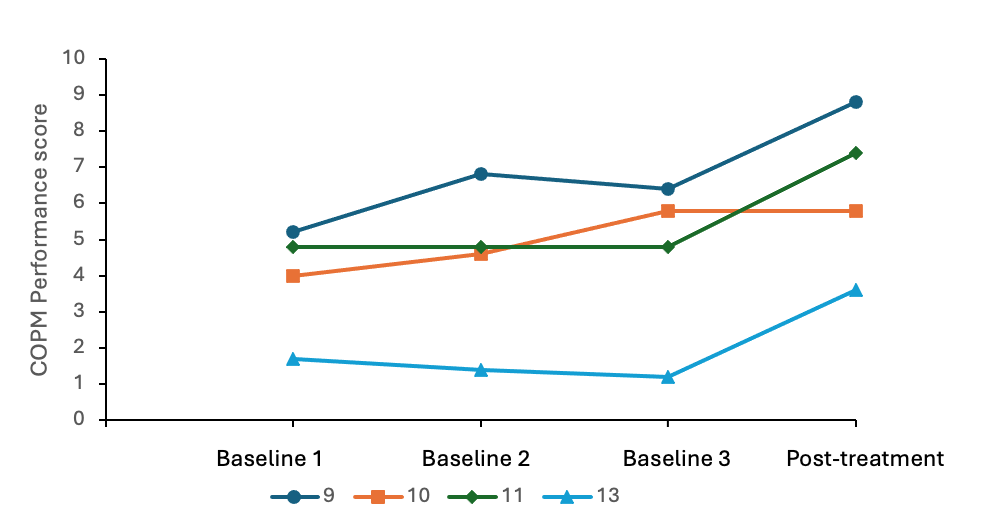

**Fig. s14. Canadian Occupational Performance Measure (COPM) Performance Scores of Treatment-As-Usual (TAU) Participants Before and After Crossover to Constraint-Induced Cognitive Therapy (CICT).** All but one showed improvement after crossover to CICT.

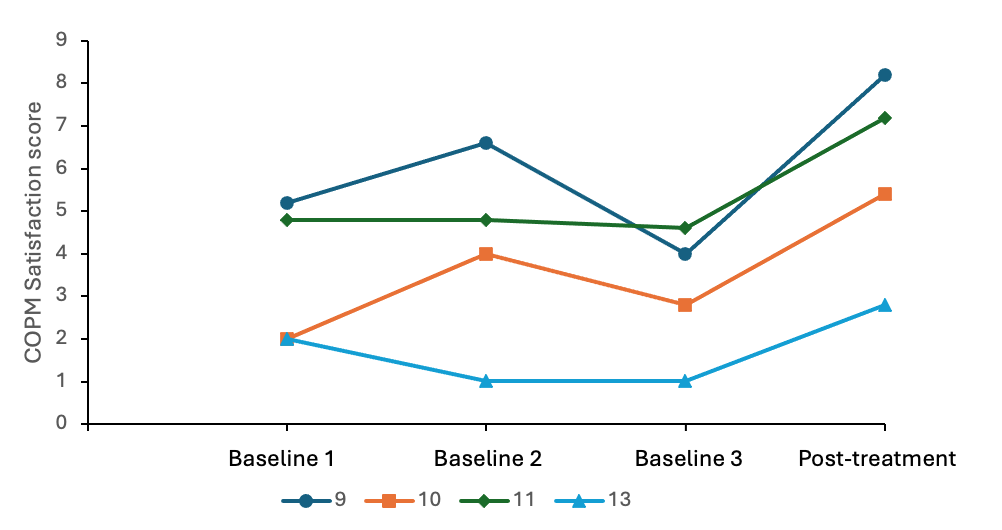

**Fig. s15. Canadian Occupational Performance Measure (COPM) Satisfaction Scores of Treatment-As-Usual (TAU) Participants Before and After Crossover to Constraint-Induced Cognitive Therapy (CICT).** All but one showed improvement after crossover to CICT.

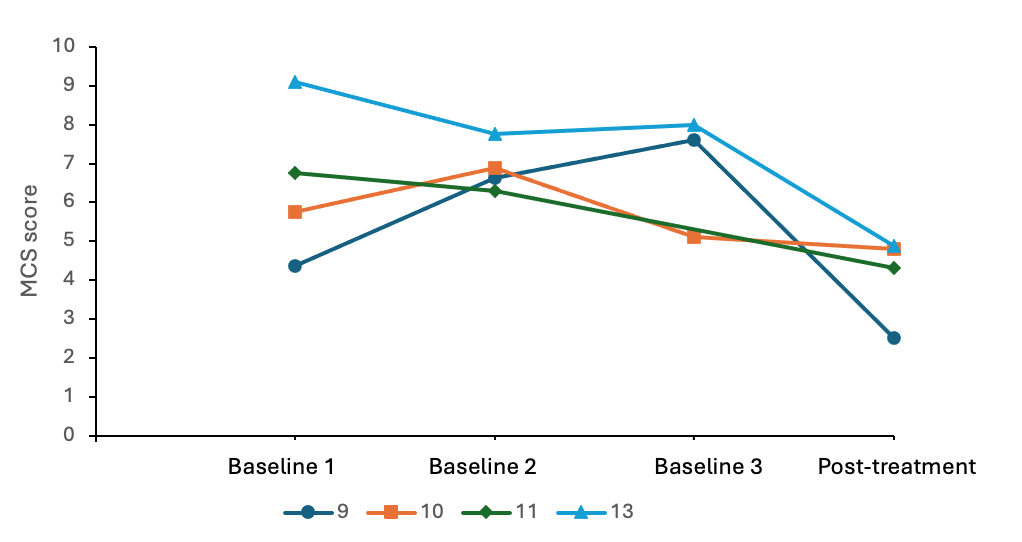

**Fig. s16. Mental Clutter Scale (MCS) Scores of Treatment-As-Usual (TAU) Participants Before and After Crossover to Constraint-Induced Cognitive Therapy (CICT).** All showed improvement after crossover to CICT.

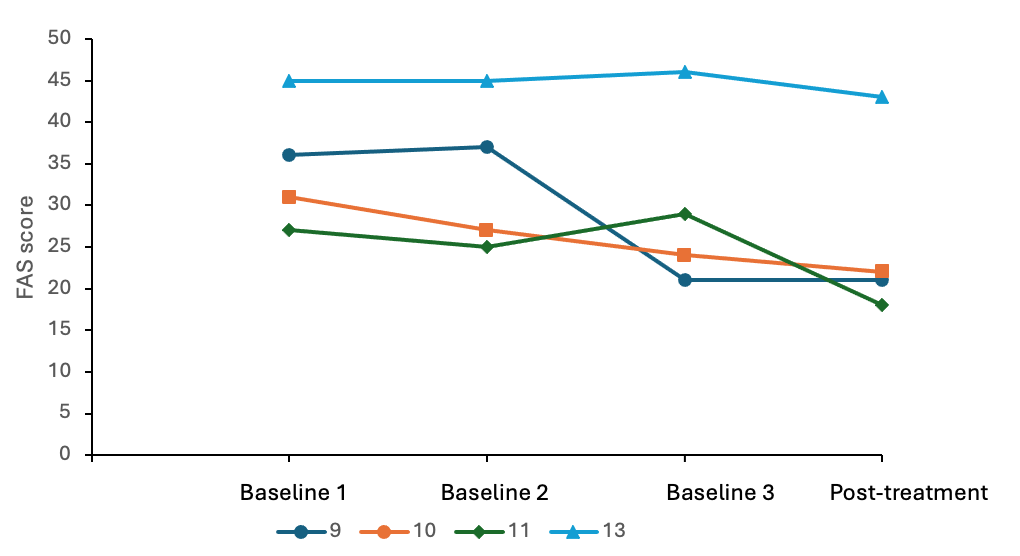

**Figure s17. Fatigue Symptom Severity (FAS) Scores of Treatment-As-Usual (TAU) Participants Before and After Crossover to CICT.** All but one showed improvement after crossover to CICT.

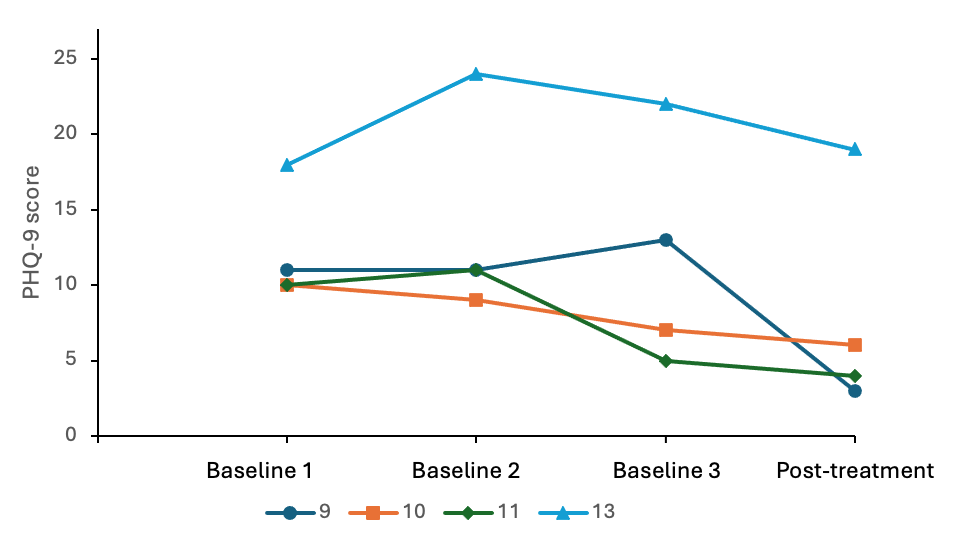

**Fig. s18. The Patient Health Questionnaire-9 (PHQ-9) Scores of Treatment-As-Usual (TAU) Participants Before and After Crossover to Constraint-Induced Cognitive Therapy (CICT).** All showed improvement after crossover to CICT.

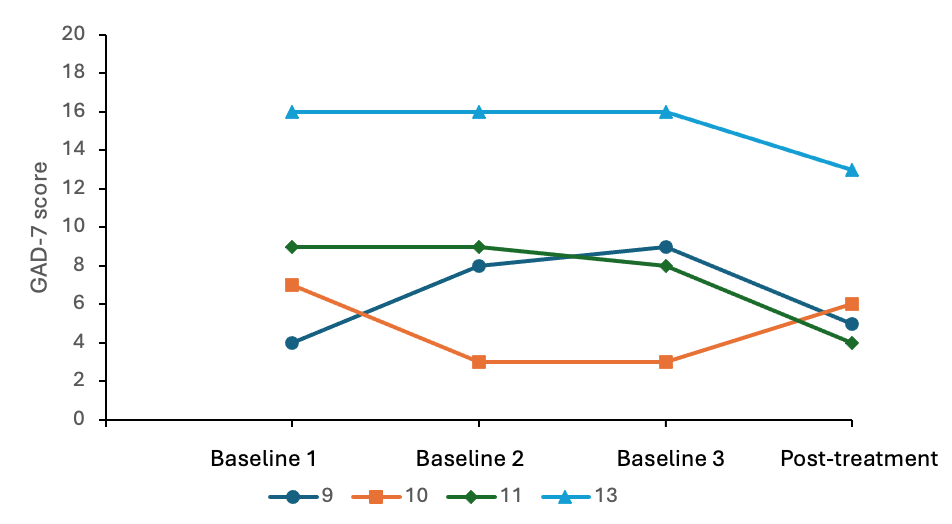

**Fig. s19. Generalized Anxiety Disorder-7 Scores of** **Treatment-As-Usual (TAU) Participants Before and After Crossover to Constraint-Induced Cognitive Therapy (CICT).** All but one showed improvement after crossover to CICT.

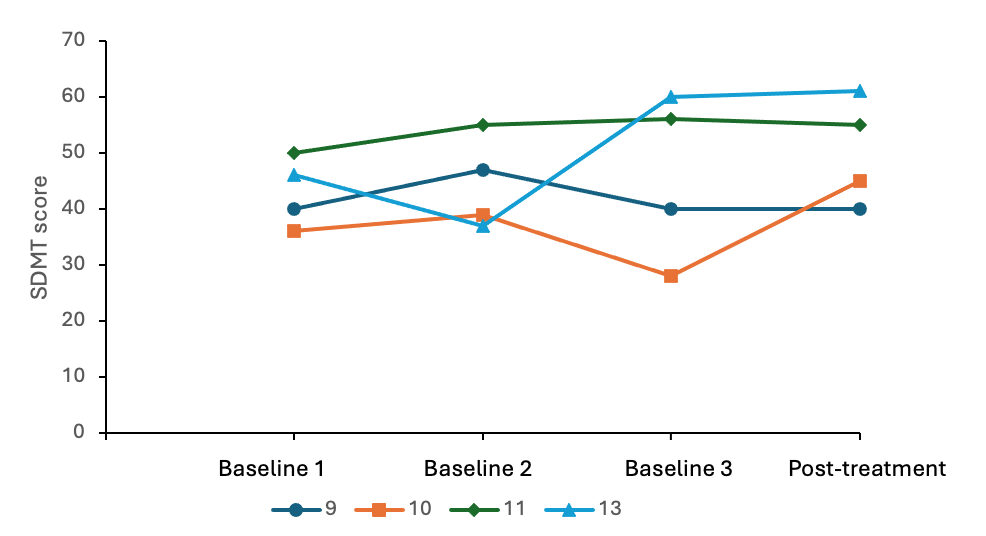

**Fig. s20. Symbol Digit Modalities Test (SDMT) Scores of Treatment-As-Usual (TAU)** **Participants Before and After Crossover to Constraint-Induced Cognitive Therapy (CICT).** Half showed improvement after crossover to CICT.

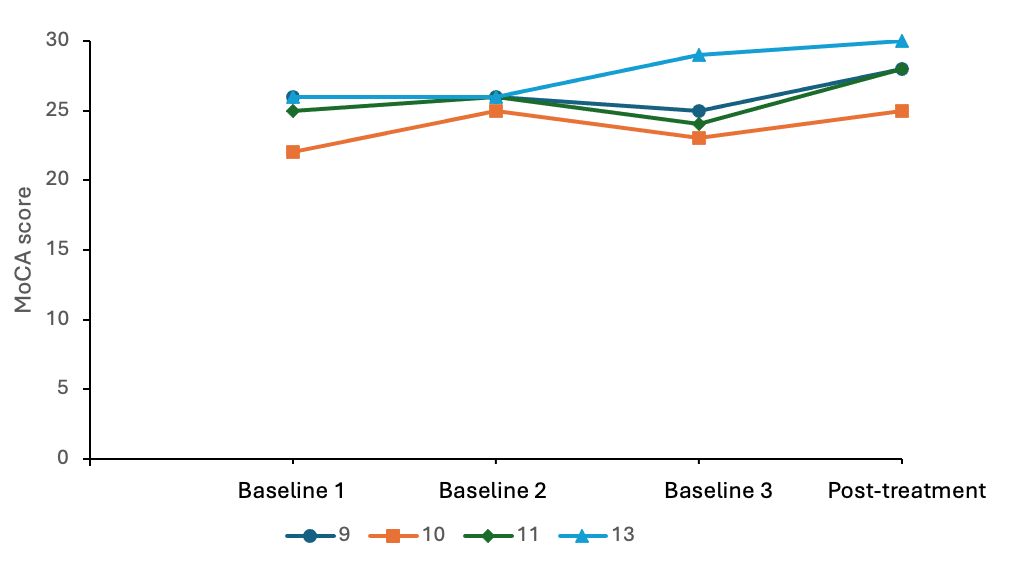

**Fig. s21. Montreal Cognitive Assessment Scores of Treatment-As-Usual (TAU) Participants Before and After Crossover to Constraint-Induced Cognitive Therapy (CICT).** All showed improvement after crossover to CICT.

**Table s7. Treatment-As-Usual (TAU) Participants’ Employment Status Before and After COVID-19 Onset and After Crossover to Constraint-Induced Cognitive Therapy (CICT)**

| Participant | Before COVID-19 | After COVID-19 | After Treatment |
| --- | --- | --- | --- |
| S9 | Media specialist | Unemployed | Returned to previous position |
| S10 | Nurse | Unemployed | Unemployed |
| S11 | Auditor | Unemployed | Started a new job |
| S13 | Tax officer | Unemployed | Unemployed |
| Summary^a^ | 100% employed | 0% employed | 50% employed |
| Note. Data for S8, S12, and S14 are not tabled because they dropped out prior to crossover to CICT. In addition, S12 and S14 were retired prior to COVID-19; return to work was not a goal for them.  ^a^ Participants were counted as employed if they could complete a full-duty set. | | | |

**DISCUSSION**

In the TAU group, outcome measure values were relatively stable across the three baseline assessments. After crossover to CICT, very large gains in performance of cognition-based activities in daily life were observed, along with very large improvements in brain fog and general cognitive ability. Large reductions were observed in fatigue and depressive and anxiety symptoms. Moderate gains were observed in cognitive processing speed. Three of four participants had clinically meaningful gains in performance of cognition-based activities in daily life and general cognitive ability. Two of the four had clinically meaningful gains in anxiety symptoms. All four TAU participants who were crossed over to CICT were unemployed before treatment. Afterwards, two of the four returned to work. The treatment response for TAU participants crossed over to CICT generally mirrored the findings from Immediate-CICT participants.

**References**

1.	Tukey JW. *Exploratory data analysis*. vol 2. Springer; 1977.

2.	Nasreddine ZS, Phillips NA, Bédirian V, et al. The Montreal Cognitive Assessment, MoCA: a brief screening tool for mild cognitive impairment. *Journal of the American Geriatrics Society*. 2005;53(4):695-699.

3.	Ball K, Edwards JD, Ross LA. The impact of speed of processing training on cognitive and everyday functions. *The Journals of Gerontology Series B: Psychological Sciences and Social Sciences*. 2007;62(Special_Issue_1):19-31.

4.	Taub E, Miller NE, Novack TA, et al. Technique to improve chronic motor deficit after stroke. *Archives of physical medicine and rehabilitation*. 1993;74(4):347-354.

5.	O'Farrell TJ, Choquette KA, Cutter H. Couples relapse prevention sessions after behavioral marital therapy for male alcoholics: outcomes during the three years after starting treatment. *Journal of studies on alcohol*. 1998;59(4):357-370.

6.	Friedman RH. Automated telephone conversations to assess health behavior and deliver behavioral interventions. *Journal of medical systems*. 1998;22:95-102.

7.	Bowers TG, Winett RA, Frederiksen LW. Nicotine fading, behavioral contracting, and extended treatment: Effects on smoking cessation. *Addictive Behaviors*. 1987;12(2):181-184.

8.	Skidmore ER, Holm MB, Whyte EM, Dew MA, Dawson D, Becker JT. The feasibility of meta-cognitive strategy training in acute inpatient stroke rehabilitation: case report. *Neuropsychological rehabilitation*. 2011;21(2):208-223.

9.	Uswatte G, Taub E, Morris D, Light K, Thompson P. The Motor Activity Log-28: assessing daily use of the hemiparetic arm after stroke. *Neurology*. 2006;67(7):1189-1194.

10.	Uswatte G, Taub E, Morris D, Vignolo M, McCulloch K. Reliability and validity of the upper-extremity Motor Activity Log-14 for measuring real-world arm use. *Stroke*. 2005;36(11):2493-2496.

11.	Haddad MM, Taub E, Uswatte G, et al. Assessing the amount of spontaneous real-world spoken language in aphasia: validation of two methods. *American Journal of Speech-Language Pathology*. 2017;26(2):316-326.

12.	Cohen J. *Statistical power analysis for the behavioral sciences*. 2nd ed. Lawrence Erlbaum Associates; 1988.
